# THE PREVALENCE OF MUSCULOSKELETAL PAIN IN AFRICA: AN OVERVIEW OF SYSTEMATIC REVIEWS WITH META-ANALYSIS INCLUDING MORE THAN 100 DISTINCT PRIMARY STUDIES

**DOI:** 10.1101/2024.05.08.24307067

**Authors:** Javier Martinez-Calderon, Marta Infante-Cano, Javier Matias-Soto, Cristina García-Muñoz

## Abstract

**Objective:** This overview of systematic reviews aimed to summarize the point, annual, and lifetime prevalence of musculoskeletal pain in African countries.

**Methods:** The CINAHL, Embase, PsycINFO, PubMed were searched until October 6, 2023. Systematic reviews with meta-analyses evaluating the prevalence of musculoskeletal pain were included. The quality of reviews was assessed with AMSTAR 2 and the overlap among reviews was calculated.

**Results:** Six reviews were included. The pooled point prevalence rate of low back pain was 39%. The pooled annual prevalence rates of low back pain ranged from 54.05% to 64.07% among meta-analyses. The pooled annual prevalence rates of upper back pain, elbow pain, wrist and/or hand pain, knee and/or leg pain, foot and/or ankle pain, and hip and/or thigh pain were 27.1%, 19.7%, 24.2%, 25.0%, 20.2%, and 15.5%, respectively. The pooled lifetime prevalence rate of low back pain was 47%. A slight overlap was found among low back pain reviews. Ethiopia, Nigeria, and South Africa were mainly studied in low back pain. The rest of types of musculoskeletal pain were only studied in Ethiopia.

**Discussion:** The prevalence of musculoskeletal pain is high. More than 100 primary studies have been meta-analyzed on this topic, underlying the high prevalence of musculoskeletal pain in African countries. Important methodological concerns were detected and discussed that can help researchers to improve and guide the future agenda in this field.

**Funding:** None.

**Review protocol:** https://doi.org/10.17605/OSF.IO/V72FY.

## Introduction

Currently, a call for action has been made to improve global health policy initiatives for preventing and treating musculoskeletal pain.(Blyth et al., 2019) During the last decade, the Analgesic, Anesthetic, and Addiction Clinical Trial Translations, Innovations, Opportunities, and Networks (ACTTION), American Pain Society (APS), and American Academy of Pain Medicine (AAPM) have developed taxonomies and diagnostic criteria for classifying acute or chronic musculoskeletal pain.(Dworkin et al., 2016; Fillingim et al., 2014; Kent et al., 2017) These taxonomies and criteria include the following diagnoses: myofascial pain (e.g., musculoskeletal low back pain), fibromyalgia, chronic widespread pain, pain-related arthritis (e.g., pain-related osteoarthritis), and temporomandibular pain disorders.

The management of musculoskeletal pain, specifically when they advance to chronicity, is challenging.(El-Tallawy et al., 2021) For example, the Global Burden of Disease Study 2017 reported low back pain is the major precursor of disability around the world.(«Global, Regional, and National Disability-Adjusted Life-Years (DALYs) for 333 Diseases and Injuries and Healthy Life Expectancy (HALE) for 195 Countries and Territories, 1990-2016: A Systematic Analysis for the Global Burden of Disease Study 2016.», 2017) The etiology and pathology of many types of chronic musculoskeletal pain (e.g., fibromyalgia) are unclear.(Ren, 2020) Cultural differences can exist when musculoskeletal pain is perceived and handled, complicating the development of protocols for managing symptoms related to this type of pain.(Reis et al., 2022) These are only some examples that characterize the complexity of musculoskeletal pain and underline the urgency of developing robust evidence-based clinical interdisciplinary approaches to mitigate the effects of musculoskeletal pain on society.

Epidemiological reviews on musculoskeletal pain help lead pain clinicians, researchers, and policymakers to better understand the burden, distribution, and determinants surrounding this type of chronic pain.(van Hecke et al., 2013) This step has thus a clear clinical relevance for deciding the prioritization and targeting of health resources. Concretely, prevalence is an epidemiological concept that could be defined in some contexts as the percentage or proportion of individuals who are experiencing a specific medical condition (e.g., musculoskeletal pain) at a given point in time or over a specified period.(*Bhopal RS. Concepts of epidemiology: integrating the ideas, theories, principles, and methods of epidemiology. Oxford: Oxford University Press; 2016.*, s. f.; *Breslow NE, Day NE, Davis W. Statistical methods in cancer research, vol. 2. Lyon: International Agency for Research on Cancer; 1987.*, s. f.; Buitrago-Garcia et al., 2022; Rothman KJ, Greenland S, Lash TL., s. f.) Oftentimes, prevalence rates are shown as point, annual, or lifetime prevalence data. Specifically in musculoskeletal pain, point prevalence can refer to the percentage or proportion of individuals reporting musculoskeletal pain at a specific point in time.(Kier, 2011) Annual prevalence can be conceptualized as the percentage or proportion of the population that has experienced musculoskeletal pain at some time during a year.(Greving et al., 2012; Jordan et al., 2007; Lassa et al., 2011) Lifetime prevalence can be defined as the percentage or proportion of people having musculoskeletal pain at some time in their life up to the moment of being interviewed.(Kessler et al., 2005)

Despite musculoskeletal pain seeming to be a clinical and research priority in countries such as England or USA,(Paskins et al., 2022) classified as high-income countries,(*The World Bank. The World by Income and Region. Available in:* https://datatopics.worldbank.org/world-development-indicators/the-world-by-income-and-region.html *Accessed [15/10/2023].*, s. f.) research efforts and funding in Africa, mainly characterized by the presence of low-income or lower-middle-income countries,(*The World Bank. The World by Income and Region. Available in:* https://datatopics.worldbank.org/world-development-indicators/the-world-by-income-and-region.html *Accessed [15/10/2023].*, s. f.) are often invested in other diseases such as HIV/AIDS, tuberculosis, or the COVID-19 pandemic.(Hartvigsen et al., 2018) However, a large number of primary evidence has been revised and meta-analyzed focusing on assessing the prevalence of musculoskeletal pain in African countries, mainly Ethiopia, Nigeria, and South Africa. (Jegnie & Afework, 2021; Mengistu et al., 2021, 2022; Morris et al., 2018) Therefore, it is time to carry out an overview of systematic reviews that synthetizes and critically analyzes the strengths and weaknesses of these reviews to show the status of musculoskeletal pain in Africa, which is essential to redefine the politics and clinical priorities to reduce the impact of musculoskeletal pain in the African population. Therefore, this overview of reviews aims to summarize and critically analyze the point, annual, and lifetime prevalence of musculoskeletal pain in African countries, based on the inclusion of systematic reviews with meta-analyses.

## Methods

We followed the Preferred Reporting Items for Overviews of Reviews (PRIOR) statement(Gates et al., 2022) and the Preferred Reporting Items for Systematic Reviews and Meta-Analyses (PRISMA) statement for abstracts.(Beller et al., 2013) We searched similar overviews of reviews in PROSPERO, the Open Science Framework (OSF), and the International Platform of Registered Systematic Review and Meta-analysis Protocols (INPLASY) to ensure that another research group was not conducting the same study. Afterward, we prospectively registered the review protocol in OSF: https://doi.org/10.17605/OSF.IO/V72FY.

### Deviations of the overview from the protocol

There are no deviations from the review protocol. *Data Sources and Search Strategy* One reviewer searched for the CINAHL (via EBSCOhost), Embase, PsycINFO (via ProQuest), and PubMed databases from inception to October 6, 2023. When possible, this reviewer used the type of document and the language of publication as search filters.

The same reviewer searched potential review articles that were not retrieved by the electronic databases in overviews of reviews or review protocols associated with the scope of this study. In addition, a manual search was conducted to search potential reviews published in French language since French is the national language for some African countries. All search strategies are reported in **Supplementary File 1**.

### Eligibility Criteria

Only articles published in peer-reviewed journals and written in English or Spanish language were included. One reviewer used the Patient, Exposure, Comparison, Outcome, Study design (PECOS) framework to carry out inclusion criteria.(Morgan et al., 2018)

### Inclusion criteria

P: People diagnosed with acute, subacute, or chronic musculoskeletal pain. We used the ACTTION-APS-AAPM taxonomies and diagnostic criteria for choosing the types of musculoskeletal pain that were included.(Dworkin et al., 2016; Fillingim et al., 2014; Kent et al., 2017) We selected the following diagnoses: myofascial pain (e.g., musculoskeletal low back pain), fibromyalgia, chronic widespread pain, pain-related arthritis (e.g., pain-related osteoarthritis), and temporomandibular pain disorders. There were no restrictions regarding age, gender, or setting.

E: Not applicable.

C: Not applicable.

O: The pooled prevalence of musculoskeletal pain. There were no restrictions regarding how the prevalence was assessed.

S: Systematic reviews with meta-analyses. Only meta-analyses reporting the prevalence data in percentages or proportions were considered. Systematic reviews were only included if they used systematic and reproducible methods to collect data on primary research studies, critically analyzed the evidence, and synthesized the results quantitatively.(Gates et al., 2022) Systematic reviews with meta-analyses should have stated the sources where the search strategies were conducted, eligibility criteria, approaches to select and extract information from primary research studies, the risk of bias assessment, and methods to analyze and summarize results that allow other researchers to replicate the findings.

### Exclusion criteria

[I] Meta-analyses or subgroup meta-analyses were not specific to musculoskeletal pain following the ACTTION-APS-AAPM taxonomies and diagnostic criteria.
[II] Meta-analyses or subgroup meta-analyses were not specific to African countries.
[III] Conference proceedings.
[IV] Impossibility of accessing full text, even after contacting via email with the corresponding author.
[V] Study protocols.

### Study Selection

One reviewer used Zotero 6.0.9 Citation Management Software to carry out the selection process. Before starting the selection process, all references were manually checked to remove duplicates.(Kwon Y, Lemieux M, McTavish J, 2015) Afterward, titles and abstracts were read, and subsequently, full texts were analyzed when abstracts seemed eligible or when abstracts were unavailable. No consensus was needed. The list with all excluded studies during the analysis of full text (n= 44) appears in **Supplementary File 2**.

### Methodological Quality Assessment

Two reviewers independently assessed the methodological quality of systematic reviews using AMSTAR 2.(Shea et al., 2017) This tool has sixteen items, seven recommended as critical domains (items: 2, 4, 7, 9, 11, 13, 15), that are rated as ‘Yes’, ‘Partially Yes’, or ‘No’. Although it is possible to use an overall score in AMSTAR 2, the authors of this tool strongly recommend not using an overall score.(Shea et al., 2017) Possible disagreements among these reviewers were solved via consensus. We calculated the percentage of agreement between these reviewers regarding the number of items rated with the same score before combining the results of their independent assessments. All the aspects related to an intervention and a comparator group did not apply to this overview since the objective is related to prevalence rates (e.g., item 1 ‘Did the research questions and inclusion criteria for the review include the components of PICO?’). The same occurred with those items whose responses are designed for systematic reviews of randomized or non-randomized studies of interventions (e.g., AMSTAR item 3 and item 9).

### Data Extraction

One reviewer extracted, whenever possible, the following analyses from each review: meta-regression analyses and/or subgroup/sensitivity meta-analyses, the methodological quality/risk of bias assessment, and the certainty of the evidence assessment using the Grading of Recommendations Assessment, Development, and Evaluation (GRADE) approach. We also aimed to extract the following information: first author and year of publication, outcome, the number of studies meta-analyzed, the total sample size meta-analyzed, pooled prevalence rates in percentages with 95% confidence intervals and I-square statistics, specific commentaries from each review (e.g., specific African countries studied). Corresponding authors were contacted via email to request or clarify some information.

### Data Synthesis

One reviewer descriptively synthesized in the main text the pooled prevalence rates of musculoskeletal pain. This reviewer grouped the results by the type of musculoskeletal pain (e.g., low back pain) and the type of prevalence parameter (e.g., lifetime prevalence). All the findings were summarized regionally (Africa) or nationally (e.g., Ethiopia), when possible. In addition, one reviewer developed maps of prevalence to depict the point, annual, or lifetime prevalence of each type of musculoskeletal pain. Maps were also developed to show the number of times that a specific musculoskeletal pain (e.g., low back pain) was studied in a specific country (e.g., Kenya) when at least two different countries were meta-analyzed in the same review. The Datawrapper app was used, developed by Datawrapper GmbH (https://app.datawrapper.de/select/map), to generate the choropleth maps. Datawrapper allowed to choose a specific African map and to show the point, annual, or lifetime prevalence rates in percentages. Some manual annotations were made once the maps were generated. When more than two reviews reported pooled prevalence rates for a specific prevalence parameter and musculoskeletal pain (e.g., annual prevalence rates for low back pain), the lowest and the highest values from meta-analyses were selected. Grey color was used in the maps to show the lack of data for some African countries.

### Overlap between Reviews

One reviewer developed a matrix of evidence to calculate the corrected covered area (CCA) for reviews evaluating low back pain. We needed to calculate the CCA to assess the overlap between systematic reviews regarding possible overlapping between primary studies.(Pieper et al., 2014) Only types of musculoskeletal pain evaluated in at least two reviews were included in the overlap calculation. The CCA refers to the area that is covered after removing the studies the first time they are counted. The overlap can be classified as slight (CCA 0-5%), moderate (CCA 6-10%), high (CCA 11-15%), or very high (CCA >15%).(Pieper et al., 2014) An upset plot was developed to depict the CCA values for low back pain.

## Results

Two hundred and sixty-three references were retrieved from the electronic databases. After removing duplicates, 134 studies were read by title and abstract, and 50 studies were analyzed in full text. Finally, six systematic reviews with meta-analyses were included.(Jegnie & Afework, 2021; Kasa et al., 2020; Mengistu et al., 2021, 2022; Morris et al., 2018; Tesfaye et al., 2023) Manually, we found four extra studies. However, these studies did not check our inclusion criteria and were excluded. Two corresponding authors were contacted to request information to decide if these studies checked our inclusion criteria.(Chikte et al., 2011; Wall et al., 2022) Both studies were excluded. The study selection process is reported in **Figure 1** and the list of excluded studies during the analysis at full text appears in **Supplementary File 2**.

**Figure 1.**
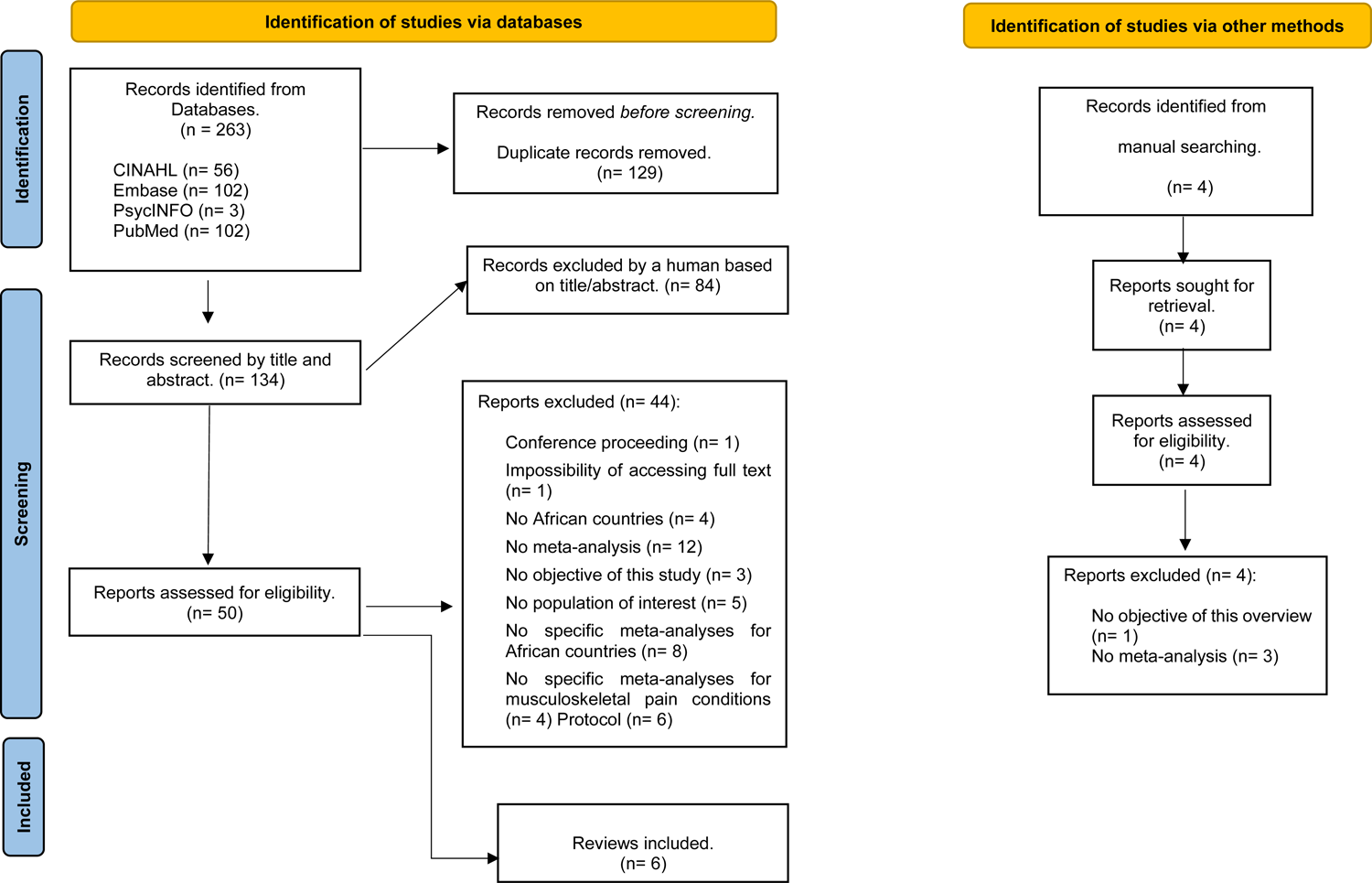
The PRISMA 2020 Flow diagram.

### Main Characteristics of Included Reviews

**Table 1** shows the characteristics of all included reviews. Most reviews were focused on low back pain.(Jegnie & Afework, 2021; Kasa et al., 2020; Mengistu et al., 2021; Morris et al., 2018; Tesfaye et al., 2023) Other types of musculoskeletal pain were studied: upper back pain,(Mengistu et al., 2021) elbow pain,(Mengistu et al., 2022) wrist and/or hand pain,(Mengistu et al., 2022) knee and/or leg pain,(Mengistu et al., 2022) foot and/or ankle pain,(Mengistu et al., 2022) as well as hip and/or thigh pain.(Mengistu et al., 2022) Pooled annual prevalence percentages were the prevalence parameter more used,(Jegnie & Afework, 2021; Kasa et al., 2020; Mengistu et al., 2021, 2022; Morris et al., 2018; Tesfaye et al., 2023) followed by point prevalence(Morris et al., 2018) and lifetime prevalence.(Morris et al., 2018) Three reviews focused on Ethiopia,(Jegnie & Afework, 2021; Mengistu et al., 2021, 2022) whereas the rest of the reviews studied different African populations.(Kasa et al., 2020; Morris et al., 2018; Tesfaye et al., 2023) These reviews mainly investigated the prevalence of musculoskeletal pain in Nigeria and South Africa (**Figure 2**). Statistically, different secondary analyses were conducted to explore potential sources of heterogeneity in meta-analyses. Three reviews used meta-regression analyses.(Jegnie & Afework, 2021; Kasa et al., 2020; Tesfaye et al., 2023) Four reviews applied sensitivity analyses.(Jegnie & Afework, 2021; Mengistu et al., 2021; Morris et al., 2018; Tesfaye et al., 2023) All reviews carried out subgroup meta-analyses. No reviews used GRADE for rating the certainty of evidence. Finally, adult and non-adult populations were evaluated, and two reviews were focused on specific populations: nurses(Kasa et al., 2020) and schoolteachers.(Tesfaye et al., 2023)

**Figure 2.**
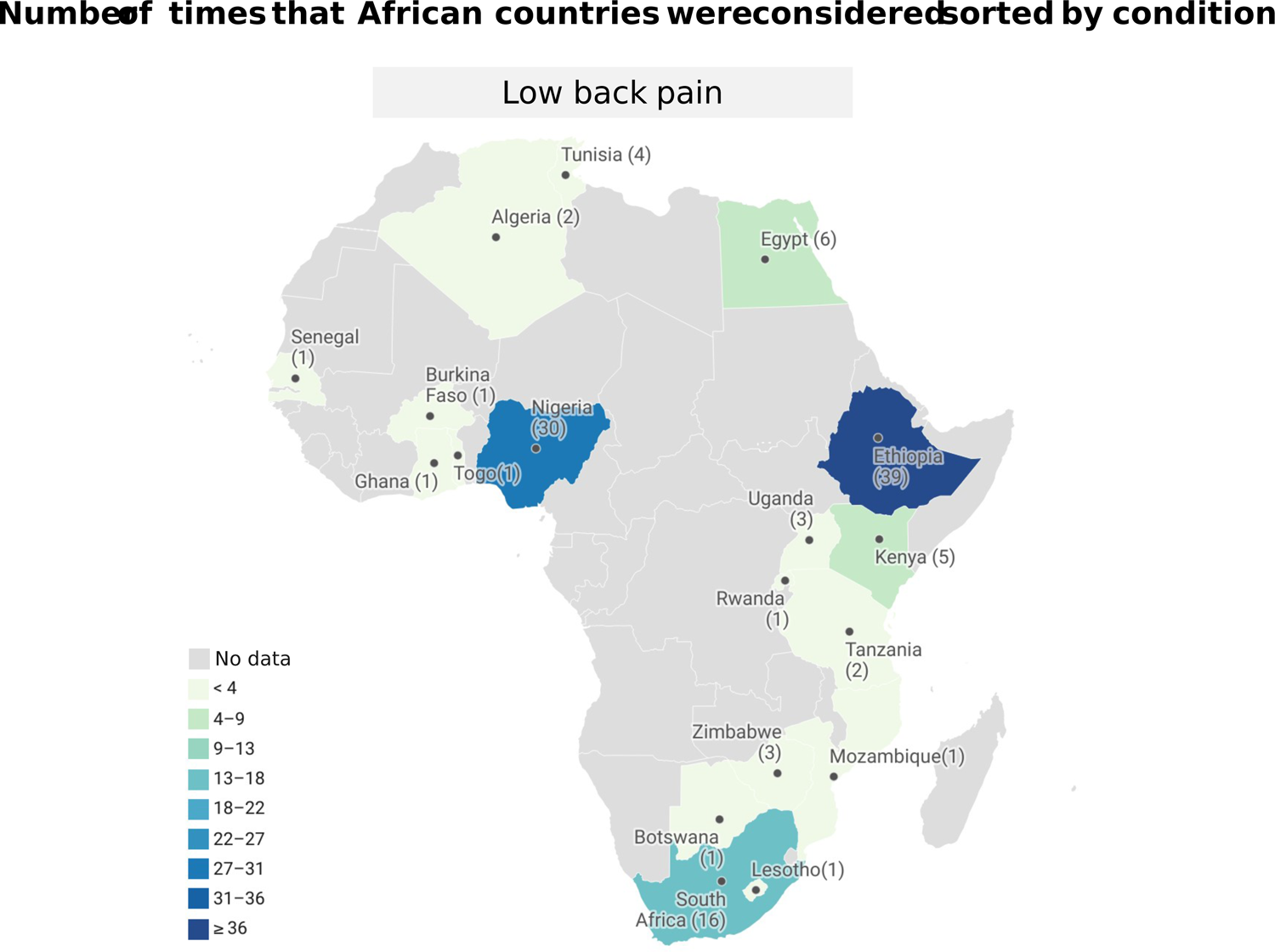
African map including the number of times a specific African country has been investigated sorted by type of musculoskeletal pain.

### Overlap between Reviews

A matrix of evidence was developed for reviews evaluating low back pain (**Supplementary File 3, Figure 3**). The was a slight overlap among reviews evaluating low back pain (N= 5, CCA= 5%).

**Figure 3.**
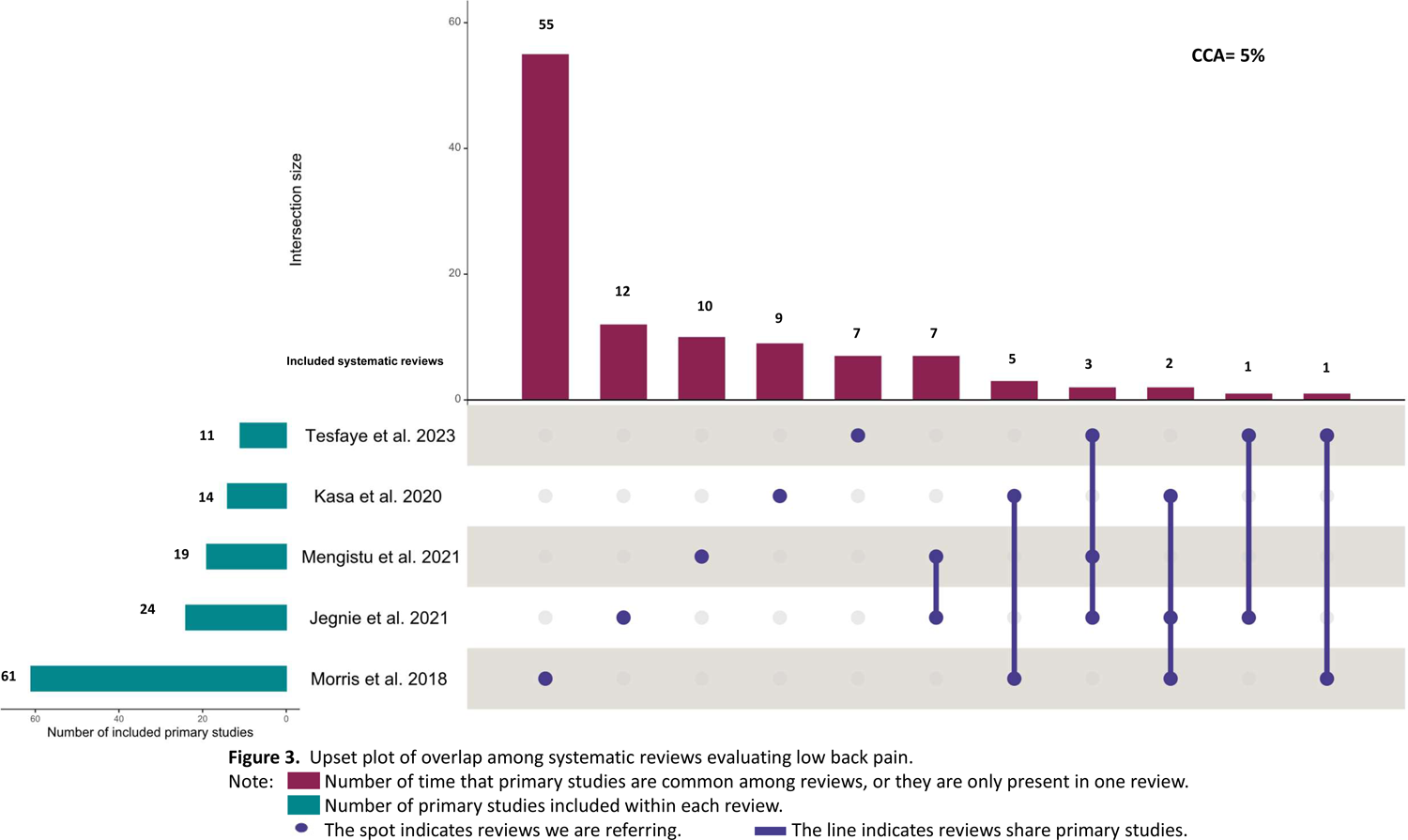
Upset plot for low back pain reviews.

**Figure 4.**
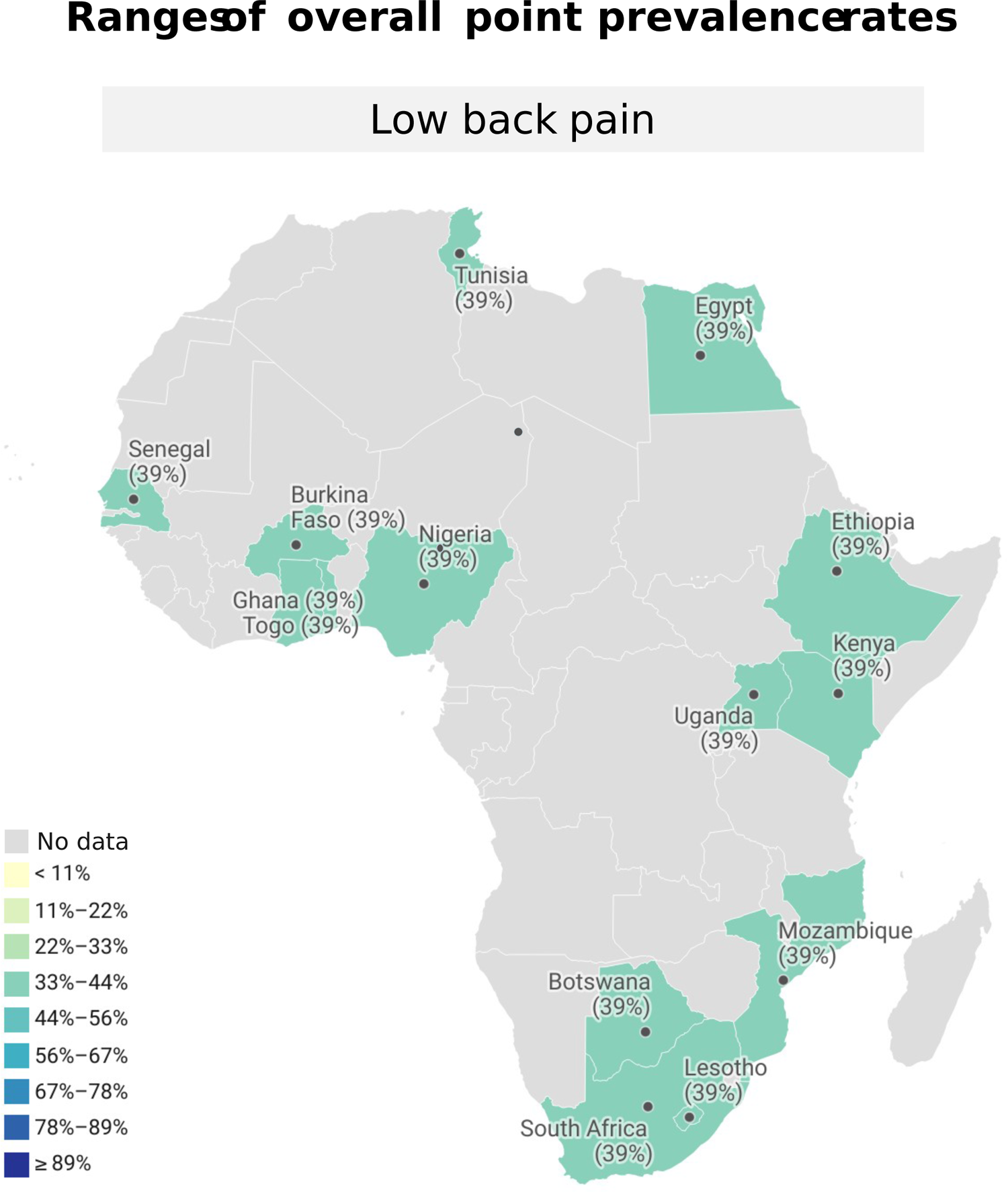
Map of point prevalence for low back pain.

**Figure 5.**
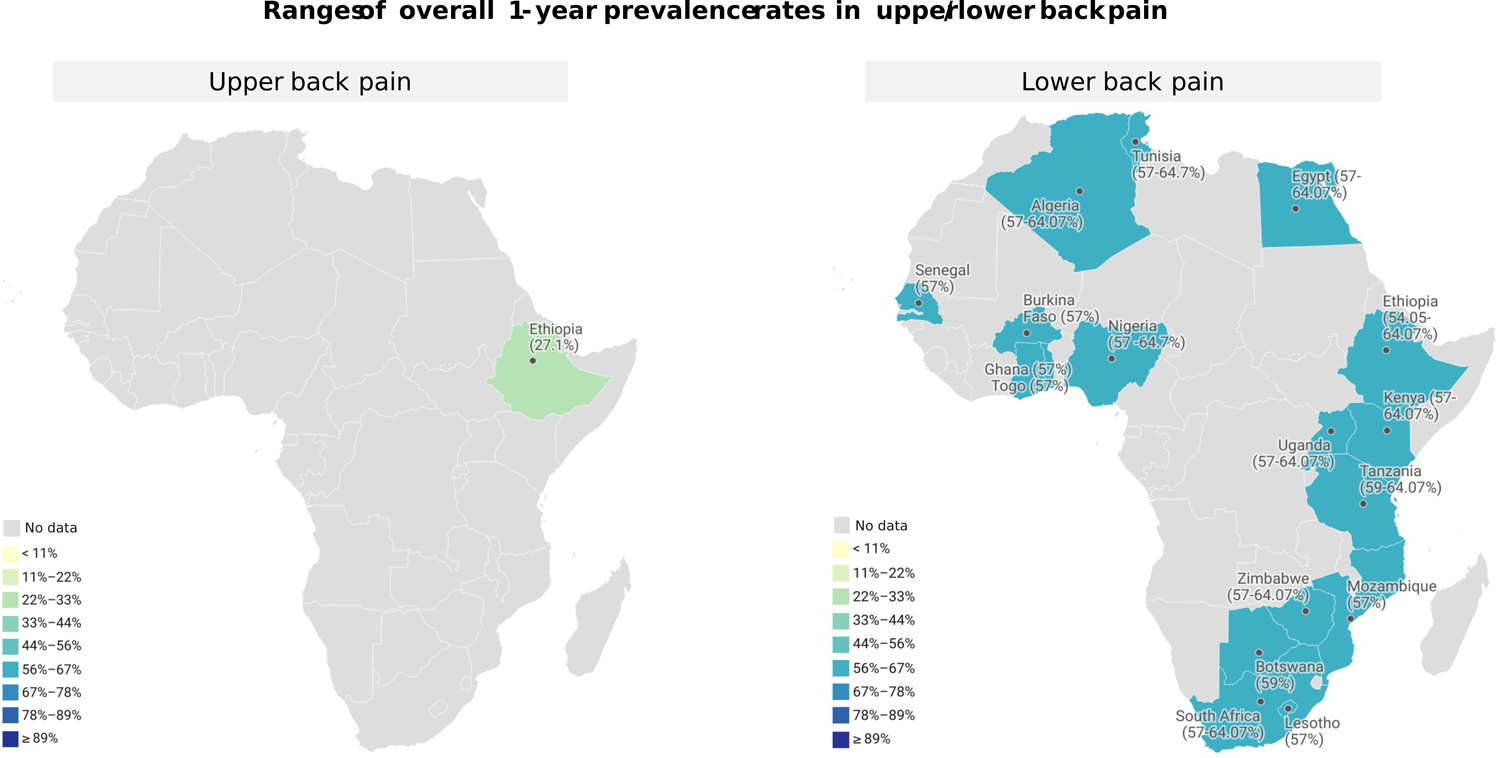
Map of annual prevalence for low back pain and upper back pain.

### Methodological Quality Assessment (AMSTAR 2)

Results from the AMSTAR 2 are described in **Table 2** (inter-rater agreement, 86.46%). Among the critical domains, none of the reviews reported or scarcely reported item 2 ‘the authors of the reviews did not prospectively register a review protocol, or they did not do so in sufficient detail’ (6/6, 100%), item 7 ‘no list of excluded studies and reasons for exclusion’ (n= 6/6, 100%), and item 4 ‘lack of a comprehensive search strategy’ (n = 6/6, 100%). Furthermore, none of the reviews reported on the sources of funding for the studies included in the review (6/6, 100%).

### Point prevalence of musculoskeletal pain

Low back pain: the pooled point prevalence of low back pain was 39% (95%CI 30-47%), I^2^= unreported (k= 23, N= 31,959) (**Figure 6**).(Morris et al., 2018)

**Figure 6.**
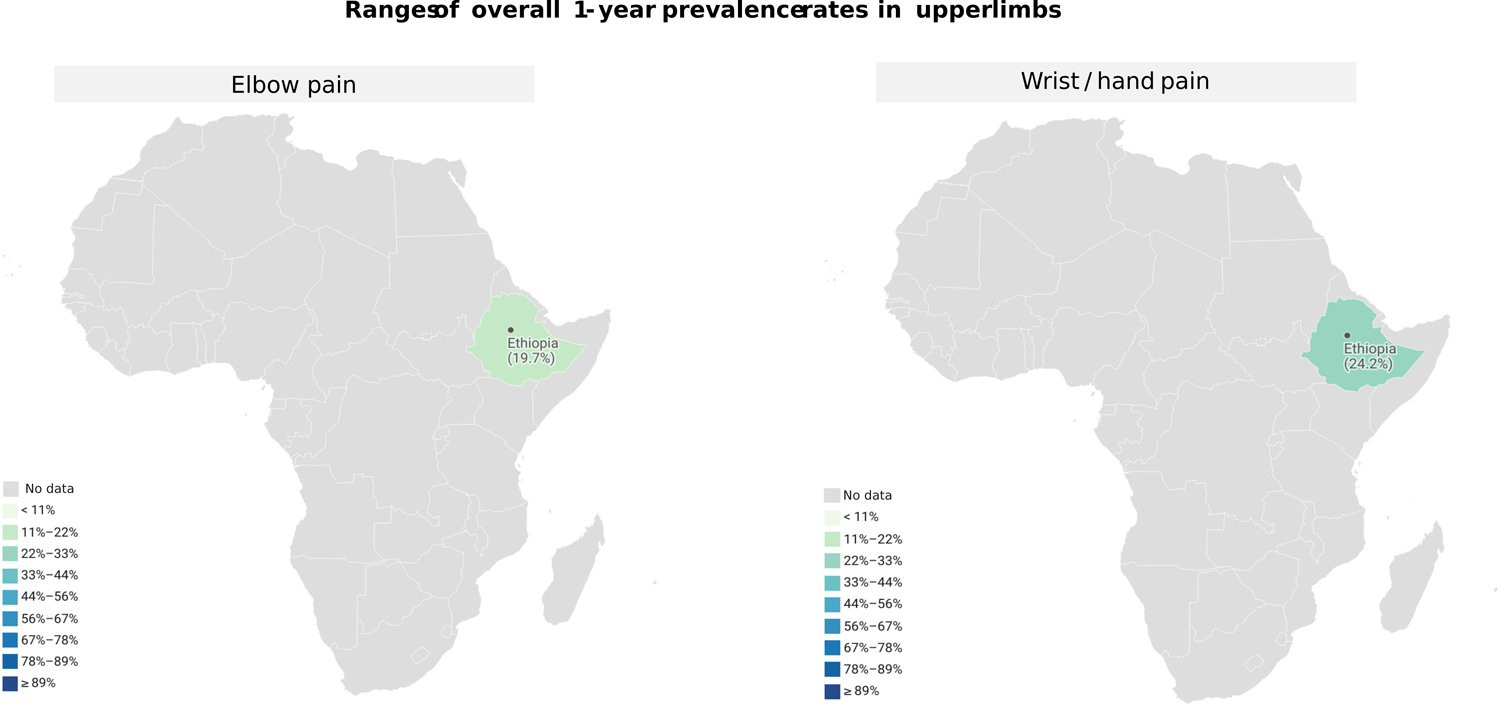
Map of annual prevalence for elbow pain and wrist and/or hand pain.

### Annual prevalence of musculoskeletal pain

Low back pain: the pooled annual prevalence of low back pain was from higher to lower prevalence percentage: 64.07% (95%CI 58.68–69.46), I^2^= 94.2% (k= 19, N= 6,110, specific population: nurses),(Kasa et al., 2020) 59.0% (95%CI 52.0-65.0%), I^2^= 96.27% (k= 11, N= 5,805, specific population: schoolteachers),(Tesfaye et al., 2023) 57% (95%CI 51-63%), I^2^= unreported (k= 34, N= 17,210),(Morris et al., 2018) 54.2% (95%CI 48.2-60.0%), I^2^= 96.78% (k= 19, N= 8,993, specific country: Ethiopia),(Mengistu et al., 2021) and 54.05% (95%CI 48.14–59.96), I^2^= 97.6% (k= 24, N= 10,447, specific country: Ethiopia) (**Figure 7**).(Jegnie & Afework, 2021) Upper back pain: the pooled annual prevalence of upper back pain was: 27.1% (95%CI 18.4-37.9), I^2^= 98.029% (k= 10, N= 4,284, specific country: Ethiopia) (**Figure 7**).(Mengistu et al., 2021) Elbow pain: the pooled annual prevalence of elbow pain was: 19.7% (95%CI 12.3-30.1%), I^2^= 98.16% (k= 10, N= 4,294, specific country: Ethiopia) (**Figure 8**).(Mengistu et al., 2022) Wrist and/or hand pain: the pooled annual prevalence of wrist and/or hand pain was: 24.2% (95%CI 17.4-32.7%), I^2^= 97.10% (k= 10, N= 4,294, specific country: Ethiopia) (**Figure 8**).(Mengistu et al., 2022) Knee and/or leg pain: the pooled annual prevalence of knee and/or leg pain was: 25.0% (18.5-32.8%), I^2^= 96.54% (k= 10, N= 4,252, specific country: Ethiopia) (**Figure 9**).(Mengistu et al., 2022) Foot and/or ankle pain: the pooled annual prevalence of foot and/or ankle pain was: 20.2% (12.8-30.4%), I^2^= 98.06% (k= 10, N= 4,252, specific country: Ethiopia) (**Figure 9**).(Mengistu et al., 2022) Hip and/or thigh pain: the pooled annual prevalence of foot and/or ankle pain was: 15.5% (9.9-23.4%), I^2^= 92.7% (k= 9, N= 3,830, specific country: Ethiopia) (**Figure 9**).(Mengistu et al., 2022)

**Figure 7.**
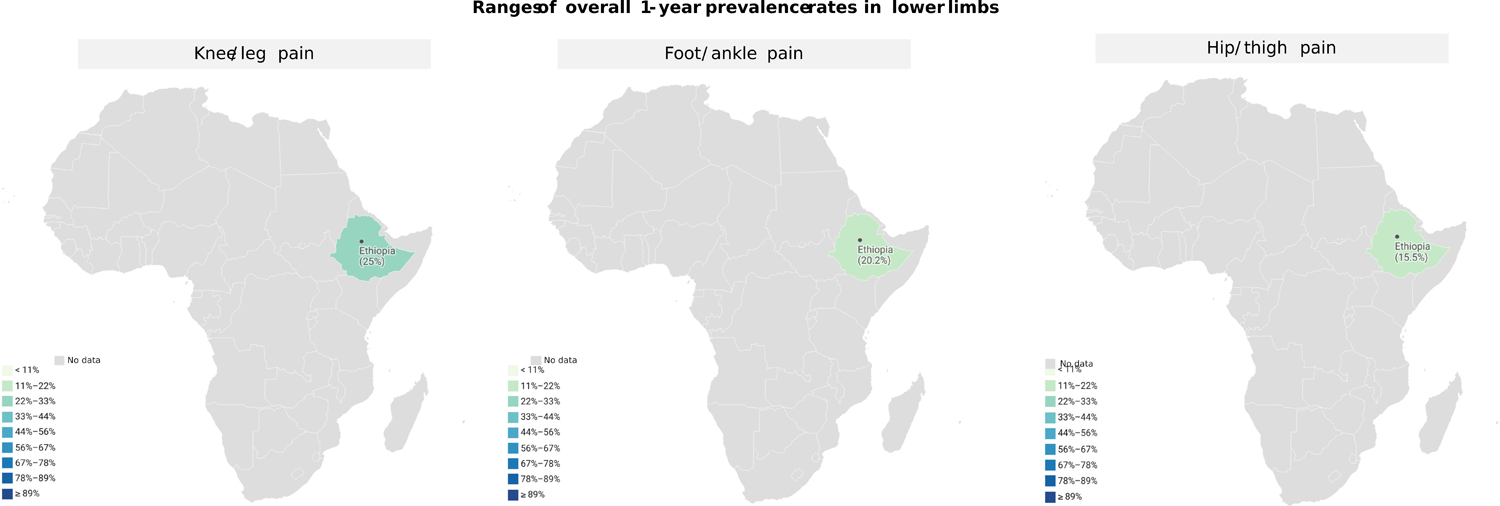
Map of annual prevalence for knee and/or leg pain, foot and/or ankle pain, and hip and/or thigh pain.

**Figure 8.**
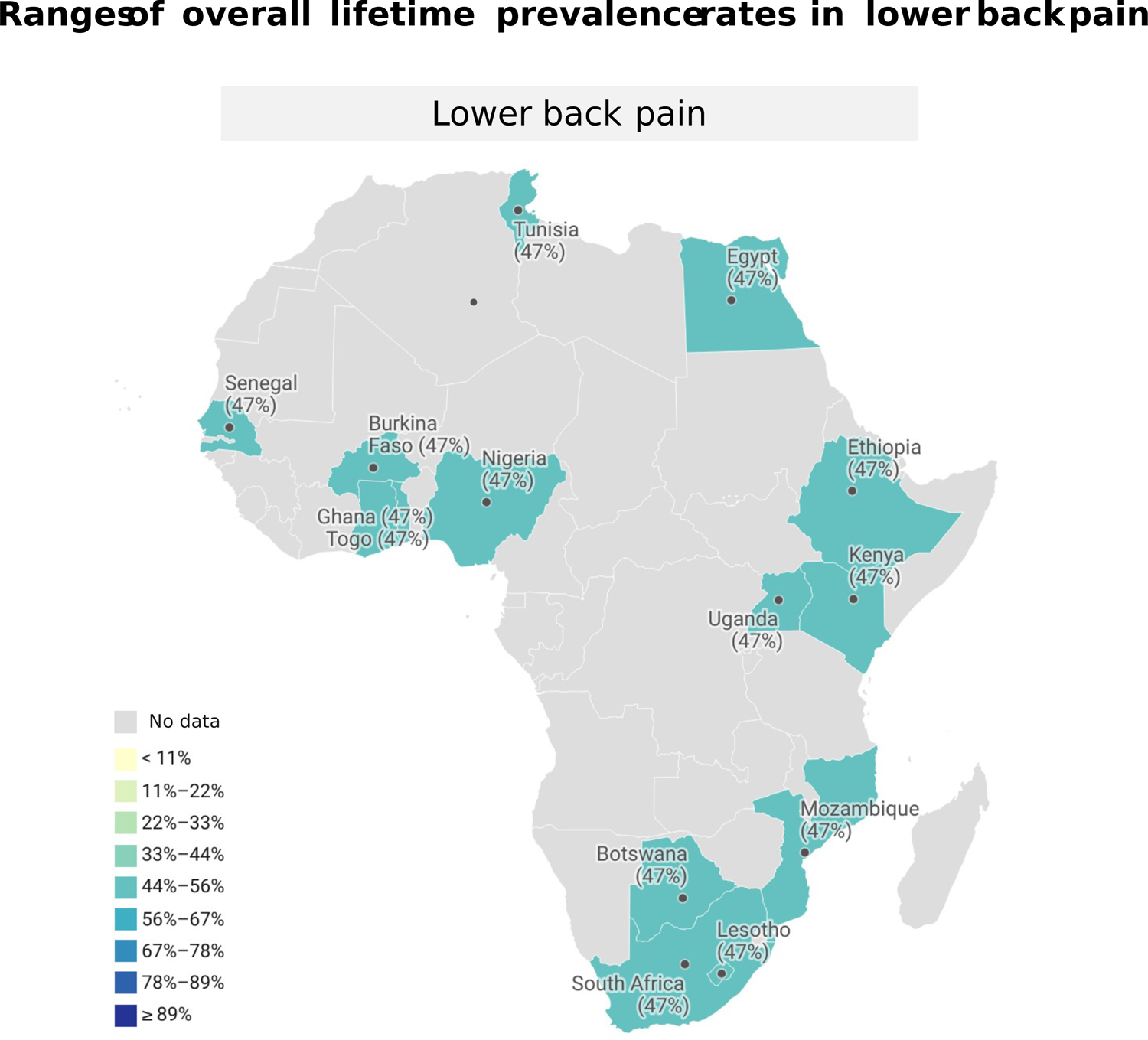
Map of lifetime prevalence for low back pain.

### Lifetime prevalence of low back pain

The pooled lifetime prevalence of low back pain was 47% (95%CI 37-58%), I^2^= unreported (k= 16, N= 14,317) (**Figure 10**).(Morris et al., 2018)

## Discussion

We aimed to synthesize the point, annual, and lifetime prevalence of musculoskeletal pain in people living in African countries, based on the inclusion of systematic reviews with meta-analyses. Overall, annual prevalence rates were the prevalence parameter more meta-analyzed. All meta-analyses showed an annual pooled prevalence superior to 15% for all musculoskeletal pain areas, underlying the pooled annual prevalence of low back pain which was superior to 50% in all meta-analyses. In addition, the pooled point or lifetime prevalence rates were also meta-analyzed in low back pain and were superior to 30% in both cases. All these pooled prevalence rates highlight musculoskeletal pain, mainly low back pain, may play an important role in African populations. We will discuss below some methodological issues can help the readers with the interpretation of the findings of this overview.

### Methodological and Clinical Considerations

One major methodological concern was related to AMSTAR-2 item 2, regarding the absence of a prospective review protocol or the lack of reporting of important details. To enhance transparency in research, clear inclusion and exclusion criteria must be provided alongside a search strategy protocol and a results analysis protocol, including assessments of bias and sources of heterogeneity. Likewise, a similar concern is observed in AMSTAR-2 item 7. Reporting the list of excluded studies along with the reasons for their exclusion would enhance the transparency of the study selection procedure of reviews. Furthermore, the search strategy must be described in detail, including clear explanations of search restrictions (e.g., temporal limitations).

This overview aimed to synthesize the prevalence of musculoskeletal pain in Africa. However, the truth is that we would be overestimating the prevalence rates that we have summarized in the results section if we conclude that these pooled findings represent the entire African continent. For example, half of the included reviews were focused on Ethiopia.(Jegnie & Afework, 2021; Mengistu et al., 2021, 2022) Less than fifteen primary studies have been meta-analyzed evaluating participants in the North of Africa (Tunisia (k= 4), Egypt (k= 6), and Algeria (k= 2)), and all of them in low back pain (**Figure 2**). Most primary studies come from Nigeria and South Africa, and non-studies have been meta-analyzed for a large list of African countries (e.g., Angola or Mauritania) (**Figure 2**). In the same vein, readers should be aware that the results would be overrepresented if we try to translate our conclusions to all types of musculoskeletal pain. Based on the ACTTION-APS-AAPM taxonomies for acute and chronic musculoskeletal pain, we made our best effort to search systematic reviews with meta-analyses analyzing the prevalence of myofascial pain, fibromyalgia, chronic widespread pain, pain-related arthritis (e.g., pain-related osteoarthritis) and temporomandibular pain disorders in African countries. However, the reality is that there is so much work to do on this continent. No meta-analyses exist for most types of musculoskeletal pain. Only low back pain(Jegnie & Afework, 2021; Kasa et al., 2020; Mengistu et al., 2021; Morris et al., 2018; Tesfaye et al., 2023) have been specifically meta-analyzed for more than one review. Furthermore, the review of Mengistu et al. 2022(Mengistu et al., 2022) combined different musculoskeletal body areas in the same meta-analysis (e.g., wrist and/or hand pain). Regarding the African population, some meta-analyses combined adult and non-adult populations, whereas other meta-analyses were focused on specific populations such as nurses(Kasa et al., 2020) or schoolteachers.(Tesfaye et al., 2023) Finally, there was a scarce number of meta-analyses regarding specific prevalence parameters. For example, the lifetime prevalence and point prevalence were only evaluated for low back pain. Only annual prevalence was considered for different musculoskeletal body areas. Therefore, we encourage readers to be critical of our results because they are preliminary and should be a call of attention to put our focus on improving the methodology of research on this topic.

No reviews used the GRADE system(Schünemann H, Brożek J, Guyatt G, Oxman A, editors., 2013) to rate the certainty of evidence. We recognize that no formal guidance has been published for using GRADE in systematic reviews of prevalence studies.(Borges Migliavaca et al., 2020) Nevertheless, we encourage systematic reviewers to use it. For example, there is information available that can help reviewers of prevalence studies to rate the GRADE dimension “risk of bias”.(Migliavaca et al., 2020) Moreover, meta-analyses and their secondary analyses allow reviewers to have specific information (e.g., statistics heterogeneity values) to rate the GRADE dimension “inconsistency”, “indirectness”, “imprecision”, and “publication bias”.

### Future research

After discussing the important gaps in knowledge that remain on this topic, we believe that the musculoskeletal pain community will benefit if more primary prevalence studies are conducted in African countries. These studies should not only consider those countries that have not been analyzed yet. But also, those African countries where we have found that the number of studies is limited (**Figure 2**). In this context, we align ourselves with prior investigations that underscore the urgent necessity for the establishment of efficient healthcare data repositories in Africa. Such repositories would facilitate the execution of rigorous studies on disease incidence, prevalence, and mortality across this continent.(Adebisi & Lucero-Prisno, 2022) Addressing this gap of knowledge would help to realize the current scale of the problem, probably reducing the disparities and underlying inequalities within African regions. Consequently, this paves the way for the strategic development of interventions aimed at preventing, promoting, and managing conditions, such as musculoskeletal pain. The same step should be also conducted for those musculoskeletal pain diagnoses (e.g., fibromyalgia or musculoskeletal shoulder pain) where prevalence rates remain unclear or are limited. In addition, we encourage pain researchers to carry out new high-quality systematic reviews with meta-analyses where they use GRADE for rating the certainty of evidence. Also, it could be essential that future high-quality systematic reviews with meta-analyses use secondary analyses to explore the differences between [I] adult and non-adult African populations, [II] African countries, [III] prevalence parameters, and [IV] musculoskeletal body areas.

### Limitations

The Cochrane Handbook recommends two reviewers should independently carry out study selection and data extraction.(Tianjing Li, Julian PT Higgins, Jonathan J Deeks, s. f.) This recommendation was not followed and the risk of biases in both methodological steps may have increased. Some important information could have been missed since we did not consider theses, dissertations, conference proceedings, systematic reviews without meta-analyses, and systematic reviews with meta-analyses published in languages other than English, French, or Spanish.

## Conclusion

The prevalence of musculoskeletal pain is high. More than 100 primary studies have been meta-analyzed on this topic, underlying the high prevalence of musculoskeletal pain in African countries. Important methodological concerns were detected and discussed that can help researchers to improve and guide the future agenda in this field.

## Ethics statement

This is a systematic review, and no ethical approval was required.

## Supporting information

Characteristics included studies

AMSTAR 2

Search strategies

Excluded studies

Overlap LBP

PRIOR checklist

PRISMA 2020 for abstracts

## Data Availability

All data produced in the present work are contained in the manuscript and supplementary files.

