## Supplementary material for "THE PREVALENCE OF MUSCULOSKELETAL PAIN IN AFRICA: AN OVERVIEW OF SYSTEMATIC REVIEWS WITH META-ANALYSIS INCLUDING MORE THAN 100 DISTINCT PRIMARY STUDIES": Characteristics included studies

**Table 1.** The characteristics of systematic reviews.

| **Study and year of publication** | **Main Analyses** | **Outcome** | **Studies (K)**  **and total of participants (N)** | **Pooled prevalence 95%CI** | **Commentary** |
| --- | --- | --- | --- | --- | --- |
| Jegnie et al. 2021(Jegnie & Afework, 2021) | Meta-regression: Yes: (1) study characteristics, (2) publication year, (3) sample size.  Sensitivity analyses: Yes: six studies were excluded from the main analyses due to methodological quality problems (from 30 studies to 24 studies).  Subgroup meta-analyses: Yes: (1) occupation of the study participants, (2) the region where the studies were conducted.  MQ assessment: the Joanna Briggs Institute Critical Appraisal Checklist for Case-Control Studies 2017 and the Newcastle Ottawa Scale (an adapted version was used for cross-sectional studies).  GRADE: No. | Presence of low back pain | k= 24, N= 10,447 | A 1-year period prevalence: 54.05% (48.14–59.96), I^2^= 97.6% | All studies were developed in Ethiopia (24).  The objective of this review was to evaluate the prevalence of occupational-related low back pain. |
| Kasa et al. 2020(Kasa et al., 2020) | Meta-regression: Yes: (1) years of the study, (2) sample size.  Sensitivity analyses: No.  Subgroup meta-analyses: Yes: (1) years of the study, (2) region in the continent (south, east, west, north).  MQ assessment: An unspecified and modified tool based on three methodological tests.  GRADE: No. | Presence of low back pain | k= 19, N= 6,110 | A 1-year period prevalence: 64.07% (58.68–69.46), I^2^= 94.2% | Studies were conducted in Algeria (1), Egypt (1), Ethiopia (4), Kenya (1), Nigeria (5), Rwanda (1), South Africa (2), Tanzania (1), Tunisia (1), Uganda (1), and Zimbabwe (1).  Adult populations (nurses) were evaluated. |
| Mengistu et al. 2021(Mengistu et al., 2021) | Meta-regression: No.  Sensitivity analyses: Yes: dropping studies that were found to influence the summary estimates.  Subgroup meta-analyses: Yes: (1) the publication year, (2) occupation categories, (3) study region, and (4) outcomes.  MQ assessment: the Joanna Briggs Institute Critical Appraisal Checklist.  GRADE: No. | Presence of upper and low back pain | k= 10, N= 4,284 | A 1-year period prevalence (previous year): upper back pain: 27.1% (18.4-37.9), I^2^= 98.029% | All studies were conducted in Ethiopia (20).  Adult populations were evaluated.  The objective of this review was to evaluate the prevalence of occupational-related low back pain. |
|  |  |  | k= 19, N= 8,993 | A 1-year period prevalence (previous year): low back pain: 54.2% (48.2-60.0%), I^2^= 96.78% |  |
| Mengistu et al. 2022(Mengistu et al., 2022) | Meta-regression: No.  Sensitivity analyses: No.  Subgroup meta-analyses: Yes: (1) the study population, (2) publication year, (3) study region where the study was conducted.  MQ assessment: the Joanna Briggs Institute Critical Appraisal Checklist for prevalence studies.  GRADE: No. | Presence of upper and or lower extremity musculoskeletal pain | k= 10, N= 4,294 | A 1-year period prevalence (previous year): elbow pain: 19.7% (12.3-30.1%), I^2^= 98.16% | All studies were conducted in Ethiopia (12).  Adult populations were evaluated.  The objective of this review was to evaluate the prevalence of occupational-related low back pain. |
|  |  |  | k= 10, N= 4,294 | A 1-year period prevalence (previous year): wrist/hand pain: 24.2% (17.4-32.7%), I^2^= 97.10% |  |
|  |  |  | k= 10, N= 4,252 | A 1-year period prevalence (previous year): knee and/or leg pain: 25.0% (18.5-32.8%), I^2^= 96.54% |  |
|  |  |  | k= 10, N= 4,252 | A 1-year period prevalence (previous year): foot and/or ankle pain: 20.2% (12.8-30.4%), I^2^= 98.06% |  |
|  |  |  | k= 9, N= 3,830 | A 1-year period prevalence (previous year): hip and/or thigh pain: 15.5% (9.9-23.4%), I^2^= 92.7% |  |
| Morris et al. 2018(Morris et al., 2018) | Meta-regression: No.  Sensitivity analyses: Yes: evaluate the presence of changes in the results in case lower methodological quality studies were included.  Subgroup meta-analyses: Yes: (1) age group (adults and children/adolescents), (2) country status (low income, low middle income, and upper middle income), (3) gender (male and female), (4) setting (community, industry, hospital, professional, and school).  MQ assessment: An unspecified tool based on 10 items, including aspects such as the definition of low back pain or the quality of data.  GRADE: No. | Presence of low back pain | k= 16, N= 14,317 (3,919 cases with low back pain) | Lifetime prevalence: 47% (37-58%), I^2^= UR | Studies were conducted in Algeria (1), Botswana (1), Burkina Faso (1), Egypt (1), Ethiopia (2), Ghana (2), Kenya (1), Lesotho (1), Mozambique (1), Nigeria (28), Senegal (1), South Africa (14), Togo (1), Tunisia (3), Uganda (2), Zimbabwe (2).  Appearing 62 studies instead of 61 because one study evaluated participants from two countries: Ethiopia and Nigeria.    Adult and non-adult populations were evaluated. |
|  |  |  | k= 34, N= 17,210 (8,627 cases with low back pain) | A 1-year period prevalence: 57% (51-63%), I^2^= UR |  |
|  |  |  | k= 23, N= 31,959 (10,454 cases with low back pain) | Point prevalence: 39% (30-47%), I^2^= UR |  |
| Tesfaye et al. 2023(Tesfaye et al., 2023) | Meta-regression: Yes: (1) age, (2) practice of physical exercise, (3) cigarette smoking, (4) presence of sleep problems, (5) history of injury.  Sensitivity analyses: Yes: excluding each study step by step.  Subgroup meta-analyses: Yes: (1) African country.  MQ assessment: the Joanna Briggs Institute Critical Appraisal Checklist for cross-sectional studies.  GRADE: No. | Presence of low back pain | k= 11, N= 5,805 | A 1-year period prevalence: 59.0% (52.0-65.0%), I^2^= 96.27% | Studies were conducted in Botswana (1), Egypt (2), Ethiopia (4), Kenya (2), Nigeria (1), and Tanzania (1).  Adult populations (schoolteachers) were evaluated. |

Note: GRADE: Grading of Recommendations Assessment, Development, and Evaluation; MQ: methodological quality; ROB: risk of bias; UR: unreported.
