## Supplementary material for "THE PREVALENCE OF MUSCULOSKELETAL PAIN IN AFRICA: AN OVERVIEW OF SYSTEMATIC REVIEWS WITH META-ANALYSIS INCLUDING MORE THAN 100 DISTINCT PRIMARY STUDIES": AMSTAR 2

**Table 2.** The methodological quality of systematic reviews (AMSTAR-2).

| **Systematic review** | **1** | **2** | **3** | **4** | **5** | **6** | **7** | **8** | **9** | **10** | **11** | **12** | **13** | **14** | **15** | **16** |
| --- | --- | --- | --- | --- | --- | --- | --- | --- | --- | --- | --- | --- | --- | --- | --- | --- |
| Jegnie et al 2021(Jegnie & Afework, 2021) |  |  |  |  |  |  |  |  |  |  |  |  |  |  |  |  |
| Kasa et al 2020(Kasa et al., 2020) |  |  |  |  |  |  |  |  |  |  |  |  |  |  |  |  |
| Mengistu et al 2021(Mengistu et al., 2021) |  |  |  |  |  |  |  |  |  |  |  |  |  |  |  |  |
| Mengistu et al 2022(Mengistu et al., 2022) |  |  |  |  |  |  |  |  |  |  |  |  |  |  |  |  |
| Morris et al 2018(Morris et al., 2018) |  |  |  |  |  |  |  |  |  |  |  |  |  |  |  |  |
| Tesfaye et al 2023(Tesfaye et al., 2023) |  |  |  |  |  |  |  |  |  |  |  |  |  |  |  |  |

**Note**: Answers: red color: No, yellow color: Partially yes, green color: Yes.

Items: AMSTAR 1: Did the research questions and inclusion criteria for the review include the components of PICO? AMSTAR 2: Did the report of the review contain an explicit statement that the review methods were established prior to the conduct of the review and did the report justify any significant deviations from the protocol? AMSTAR 3: Did the review authors explain their election of the study designs for inclusion in the review? AMSTAR 4: Did the review authors use a comprehensive literature search strategy? AMSTAR 5: Did the review authors perform study selection in duplicate? AMSTAR 6: Did the review authors perform data extraction in duplicate? AMSTAR 7: Did the review authors provide a list of excluded studies and justify the exclusions? AMSTAR 8: Did the review authors describe the included studies in adequate detail? AMSTAR 9: Did the review authors use a satisfactory technique for assessing the risk of bias in individual studies that were included in the review? AMSTAR 10: Did the review authors report on the sources of funding for the studies included in the review? AMSTAR 11: If meta-analysis was performed did the review authors use appropriate methods for statistical combination of results? AMSTAR 12: If meta-analysis was performed, did the review authors assess the potential impact of risk of bias in individual studies on the results of the meta-analysis or other evidence synthesis? AMSTAR 13: Did the review authors account for risk of bias in individual studies when interpreting/ discussing the results of the review? AMSTAR 14: Did the review authors provide a satisfactory explanation for, and discussion of, any heterogeneity observed in the results of the review? AMSTAR 15: If they performed quantitative synthesis did the review authors carry out an adequate investigation of publication bias (small study bias) and discuss its likely impact on the results of the review? AMSTAR 16: Did the review authors report any potential sources of conflict of interest, including any funding they received for conducting the review?
