## Supplementary material for "THE PREVALENCE OF MUSCULOSKELETAL PAIN IN AFRICA: AN OVERVIEW OF SYSTEMATIC REVIEWS WITH META-ANALYSIS INCLUDING MORE THAN 100 DISTINCT PRIMARY STUDIES": Search strategies

**Supplementary file 1.** Search Strategies**.**

**CINAHL (date 06/10/2023)**

TI (Pain OR neck OR cervical OR whiplash OR shoulder OR rotator-cuff OR back OR low-back OR thoracic OR chest OR elbow OR epicondylalgia OR wrist OR hand OR hip OR knee OR osteoarthritis OR arthrosis OR arthritis OR ankle OR musculoskeletal OR myofascial OR muscle OR pelvic OR groin OR ankylosing OR spondyl* OR gout OR fibromyalgia OR temporomandibular) AND AB (prevalence OR prevalent) AND TI (systematic-review OR meta-analysis OR meta-review OR meta-analytic-review OR metaanalysis OR meta-analyses) AND TX (Africa OR African OR Algeria OR Ethiopia OR Nigeria OR Angola OR Gabon OR Rwanda OR Benin OR Gambia OR Sao-Tome-and-Principe OR Botswana OR Ghana OR Burkina-Faso OR Guinea OR Burundi OR Senegal OR Cabo-Verde OR Kenya OR Seychelles OR Cameroon OR Lesotho OR Sierra-Leone OR Central-African-Republic OR Liberia OR South-Africa OR Chad OR Madagascar OR South-Sudan OR Comoros OR Malawi OR Togo OR Congo OR Mali OR Uganda OR Ivoire OR Mauritania OR Tanzania OR Congo OR Mauritius OR Zambia OR Mozambique OR Zimbabwe OR Eritrea OR Namibia OR Eswatini OR Niger OR Morocco OR Tunisia OR Libya OR Egypt OR Somalia)

Search modes - Boolean/Phrase.

Search filter: publication type: academic publications AND journals.

Search filter: language: English OR Spanish.

**Studies retrieved = 56**

**Embase (date 06/10/2023)**

(pain:ti OR neck:ti OR cervical:ti OR whiplash:ti OR shoulder:ti OR 'rotator cuff':ti OR back:ti OR 'low back':ti OR thoracic:ti OR chest:ti OR elbow:ti OR epicondylalgia:ti OR wrist:ti OR hand:ti OR hip:ti OR knee:ti OR osteoarthritis:ti OR arthrosis:ti OR arthritis:ti OR ankle:ti OR musculoskeletal:ti OR myofascial:ti OR muscle:ti OR pelvic:ti OR groin:ti OR ankylosing:ti OR spondyl*:ti OR gout:ti OR fibromyalgia:ti OR temporomandibular:ti) AND (prevalence:ab,ti OR prevalent:ab,ti) AND ('systematic review':ti OR 'meta analysis':ti OR 'meta review':ti OR 'meta analytic review':ti OR metaanalysis:ti OR 'meta analyses':ti) AND (africa OR african OR algeria OR ethiopia OR nigeria OR angola OR gabon OR rwanda OR benin OR gambia OR 'sao tome and principe' OR botswana OR ghana OR 'burkina faso' OR guinea OR burundi OR senegal OR 'cabo verde' OR kenya OR seychelles OR cameroon OR lesotho OR 'sierra leone' OR 'central african republic' OR liberia OR 'south africa' OR chad OR madagascar OR 'south sudan' OR comoros OR malawi OR togo OR mali OR uganda OR ivoire OR mauritania OR tanzania OR congo OR mauritius OR zambia OR mozambique OR zimbabwe OR eritrea OR namibia OR eswatini OR niger OR morocco OR tunisia OR libya OR egypt OR somalia)

Search filter: publication type (review OR article OR preprint).

Search filter: language: English OR Spanish.

**Studies retrieved = 102**

**PsycINFO (date 06/10/2023)**

title((Pain OR neck OR cervical OR whiplash OR shoulder OR rotator-cuff OR back OR low back OR thoracic OR chest OR elbow OR epicondylalgia OR wrist OR hand OR hip OR knee OR osteoarthritis OR arthrosis OR arthritis OR ankle OR musculoskeletal OR myofascial OR muscle OR pelvic OR groin OR ankylosing OR spondyl* OR gout OR fibromyalgia OR temporomandibular) ) AND abstract((prevalence OR prevalent) ) AND title((systematic-review OR meta-analysis OR meta-review OR meta-analytic-review OR metaanalysis OR meta-analyses) ) AND (Africa OR African OR Algeria OR Ethiopia OR Nigeria OR Angola OR Gabon OR Rwanda OR Benin OR Gambia OR Sao-Tome-and-Principe OR Botswana OR Ghana OR Burkina-Faso OR Guinea OR Burundi OR Senegal OR Cabo-Verde OR Kenya OR Seychelles OR Cameroon OR Lesotho OR Sierra-Leone OR Central-African-Republic OR Liberia OR South-Africa OR Chad OR Madagascar OR Sudan OR Comoros OR Malawi OR Togo OR Congo OR Mali OR Uganda OR Ivoire OR Mauritania OR Tanzania OR Congo OR Mauritius OR Zambia OR Mozambique OR Zimbabwe OR Eritrea OR Namibia OR Eswatini OR Niger OR Morocco OR Tunisia OR Libya OR Egypt OR Somalia)

Search modes - Boolean/Phrase.

Search filter: publication type: academic publications AND journals.

Search filter: language: English OR Spanish.

**Studies retrieved = 3**

**PubMed (date 06/10/2023)**

(Pain [title] OR neck [title] OR cervical [title] OR whiplash [title] OR shoulder [title] OR rotator-cuff [title] OR back [title] OR low-back [title] OR thoracic [title] OR chest [title] OR elbow [title] OR epicondylalgia [title] OR wrist [title] OR hand [title] OR hip [title] OR knee [title] OR osteoarthritis [title] OR arthrosis [title] OR arthritis [title] OR ankle [title] OR musculoskeletal [title] OR myofascial [title] OR muscle [title] OR pelvic [title] OR groin [title] OR ankylosing [title] OR spondyl* [title] OR gout [title] OR fibromyalgia [title] OR temporomandibular [title]) AND (prevalence [tiab] OR prevalent [tiab]) AND (systematic-review [title] OR meta-analysis [title] OR meta-review [title] OR meta-analytic-review [title] OR metaanalysis [title] OR meta-analyses [title]) AND (Africa [all] OR African [all] OR Algeria [all] OR Ethiopia [all] OR Nigeria [all] OR Angola [all] OR Gabon [all] OR Rwanda [all] OR Benin [all] OR Gambia [all] OR Sao-Tome-and-Principe [all] OR Botswana [all] OR Ghana [all] OR Burkina-Faso [all] OR Guinea [all] OR Burundi [all] OR Senegal [all] OR Cabo-Verde [all] OR Kenya [all] OR Seychelles [all] OR Cameroon [all] OR Lesotho [all] OR Sierra-Leone [all] OR Central-African-Republic [all] OR Liberia [all] OR South-Africa [all] OR Chad [all] OR Madagascar [all] OR South-Sudan [all] OR Comoros [all] OR Malawi [all] OR Togo [all] OR Congo [all] OR Mali [all] OR Uganda [all] OR Ivoire [all] OR Mauritania [all] OR Tanzania [all] OR Congo [all] OR Mauritius [all] OR Zambia [all] OR Mozambique [all] OR Zimbabwe [all] OR Eritrea [all] OR Namibia [all] OR Eswatini [all] OR Niger [all] OR Morocco [all] OR Tunisia [all] OR Libya [all] OR Egypt [all] OR Somalia [all])

**Studies retrieved = 102**

**Manual search**

**Review articles published in French language using the search strategies described above.**

CINAHL = 0.

Embase = one article retrived but was irrelevant for the scope of this overview.

Hannachi H, Chelly S, Ben Hassine D, Chebil D, Melki S, Nouira S, Merzougui L, Ben Abdelaziz A. Effectiveness of hand hygiene in an epidemic context. Systematic review. Tunis Med. 2020 Nov;98(11):763-771.

PsycINFO = 0.

PubMed = 0.
