## Supplementary material for "THE PREVALENCE OF MUSCULOSKELETAL PAIN IN AFRICA: AN OVERVIEW OF SYSTEMATIC REVIEWS WITH META-ANALYSIS INCLUDING MORE THAN 100 DISTINCT PRIMARY STUDIES": Excluded studies

**Supplementary file 2.** List of excluded studies that were reviewed in full text (n= 44): specific reasons.

- Conference proceeding (n= 1)
- Impossibility of accessing full text (n= 1)
- No African countries (n= 4)
- No meta-analysis (n= 12)
- No objective of this study (n= 3)
- No population of interest (n= 5)
- No specific meta-analyses for African countries (n= 8)
- No specific meta-analyses for musculoskeletal pain conditions (n= 4)
- Protocol (n= 6)

|  | **Excluded reviews** | **Reason** |
| --- | --- | --- |
| 1. | Abdullahi A, Candan SA, Abba MA, Bello AH, Alshehri MA, Afamefuna Victor E, Umar NA, Kundakci B. Neurological and Musculoskeletal Features of COVID-19: A Systematic Review and Meta-Analysis. Front Neurol. 2020 Jun 26;11:687. doi: 10.3389/fneur.2020.00687. | No objective of this study.  Despite this review included information about the prevalence of musculoskeletal pain (e.g., back pain), the aim of this review was to summarize the evidence on the neurological and musculoskeletal symptoms of COVID-19 pandemic. |
| 2. | Alinaghi F, Calov M, Kristensen LE, Gladman DD, Coates LC, Jullien D, Gottlieb AB, Gisondi P, Wu JJ, Thyssen JP, Egeberg A. Prevalence of psoriatic arthritis in patients with psoriasis: A systematic review and meta-analysis of observational and clinical studies. J Am Acad Dermatol. 2019 Jan;80(1):251-265.e19. doi: 10.1016/j.jaad.2018.06.027. | No population of interest. |
| 3. | Allado E, Moussu A, Bigna JJ, Hamroun S, Camier A, Chenuel B, Hamroun A. Global prevalence of spondyloarthritis in low-income and middle-income countries: a systematic review and meta-analysis protocol. BMJ Open. 2020 Oct 29;10(10):e041180. doi: 10.1136/bmjopen-2020-041180. | Protocol. |
| 4. | Almutairi K, Nossent J, Preen D, Keen H, Inderjeeth C. The global prevalence of rheumatoid arthritis: a meta-analysis based on a systematic review. Rheumatol Int. 2021 May;41(5):863-877. doi: 10.1007/s00296-020-04731-0. | No population of interest. |
| 5. | Ardakani EM, Leboeuf-Yde C, Walker BF. Failure to define low back pain as a disease or an episode renders research on causality unsuitable: results of a systematic review. Chiropr Man Therap. 2018 Jan 9;26:1. doi: 10.1186/s12998-017-0172-9. | No meta-analysis. |
| 6. | Asante BO, Trask C, Adebayo O, Bath B. Prevalence and risk factors of low back disorders among waste collection workers: A systematic review. Work. 2019;64(1):33-42. doi: 10.3233/WOR-192977. | No meta-analysis. |
| 7. | Ayouni I, Chebbi R, Hela Z, Dhidah M. Comorbidity between fibromyalgia and temporomandibular disorders: a systematic review. Oral Surg Oral Med Oral Pathol Oral Radiol. 2019 Jul;128(1):33-42. doi: 10.1016/j.oooo.2019.02.023. | No meta-analysis. |
| 8. | Barake M, El Eid R, Ajjour S, Chakhtoura M, Meho L, Mahmoud T, Atieh J, Sibai AM, El-Hajj Fuleihan G. Osteoporotic hip and vertebral fractures in the Arab region: a systematic review. Osteoporos Int. 2021 Aug;32(8):1499-1515. doi: 10.1007/s00198-021-05937-z. | No meta-analysis. |
| 9. | Bernal D, Campos-Serna J, Tobias A, Vargas-Prada S, Benavides FG, Serra C. Work-related psychosocial risk factors and musculoskeletal disorders in hospital nurses and nursing aides: a systematic review and meta-analysis. Int J Nurs Stud. 2015 Feb;52(2):635-48. doi: 10.1016/j.ijnurstu.2014.11.003. | No specific meta-analyses for African countries.  This review only include one study evaluating one African country. |
| 10. | Chikte UM, Khondowe O, Louw Q, Musekiwa A. A meta analysis of the prevalence of spinal pain among dentists. SADJ. 2011 Jun;66(5):214-8. | Impossibility of accessing full text.  We tried to contact with the authors by we did not receive any response. |
| 11. | Essouma M, Nkeck JR, Endomba FT, Bigna JJ, Ralandison S. Epidemiology of rheumatoid arthritis in sub-Saharan Africa: a systematic review and meta-analysis protocol. Syst Rev. 2020 Apr 17;9(1):81. doi: 10.1186/s13643-020-01342-5. | Protocol. |
| 12. | Fan Z, Yan L, Liu H, Li X, Fan K, Liu Q, Li JJ, Wang B. The prevalence of hip osteoarthritis: a systematic review and meta-analysis. Arthritis Res Ther. 2023 Mar 29;25(1):51. doi: 10.1186/s13075-023-03033-7. | No specific meta-analyses for African countries.  This review only include one study evaluating one African country. |
| 13. | Fernández-de-Las-Peñas C, Navarro-Santana M, Plaza-Manzano G, Palacios-Ceña D, Arendt-Nielsen L. Time course prevalence of post-COVID pain symptoms of musculoskeletal origin in patients who had survived severe acute respiratory syndrome coronavirus 2 infection: a systematic review and meta-analysis. Pain. 2022 Jul 1;163(7):1220-1231. doi: 10.1097/j.pain.0000000000002496. | No specific meta-analyses for African countries.  This review only include one study evaluating one African country: Egypt. |
| 14. | Gebreyesus T, Nigussie K, Gashaw M, Janakiraman B. The prevalence and risk factors of work-related musculoskeletal disorders among adults in Ethiopia: a study protocol for extending a systematic review with meta-analysis of observational studies. Syst Rev. 2020 Jun 8;9(1):136. doi: 10.1186/s13643-020-01403-9. | Protocol. |
| 15. | Gebrye T, Niyi-Odumosu F, Lawoe J, Mbada C, Fatoye F. The impact of COVID-19 related lockdown restrictions on musculoskeletal health: a systematic review. Rheumatol Int. 2023 Nov;43(11):2011-2019. doi: 10.1007/s00296-023-05406-2. | No meta-analysis. |
| 16. | Harle CA, Danielson EC, Derman W, Stuart M, Dvorak J, Smith L, Hainline B. Analgesic Management of Pain in Elite Athletes: A Systematic Review. Clin J Sport Med. 2018 Sep;28(5):417-426. doi: 10.1097/JSM.0000000000000604. | No meta-analysis. |
| 17. | Hill J, Harrison J, Christian D, Reed J, Clegg A, Duffield SJ, Goodson N, Marson T. The prevalence of comorbidity in rheumatoid arthritis: a systematic review and meta-analysis. Br J Community Nurs. 2022 May 2;27(5):232-241. doi: 10.12968/bjcn.2022.27.5.232. | No specific meta-analyses for African countries. |
| 18. | Jackson T, Thomas S, Stabile V, Han X, Shotwell M, McQueen K. Prevalence of chronic pain in low-income and middle-income countries: a systematic review and meta-analysis. Lancet. 2015 Apr 27;385 Suppl 2:S10. doi: 10.1016/S0140-6736(15)60805-4. | Conference proceeding. |
| 19. | Jacquier-Bret J, Gorce P. Prevalence of Body Area Work-Related Musculoskeletal Disorders among Healthcare Professionals: A Systematic Review. Int J Environ Res Public Health. 2023 Jan 2;20(1):841. doi: 10.3390/ijerph20010841. | No meta-analysis. |
| 20. | Kaka B, Maharaj SS, Fatoye F. Prevalence of musculoskeletal disorders in patients with diabetes mellitus: A systematic review and meta-analysis. J Back Musculoskelet Rehabil. 2019;32(2):223-235. doi: 10.3233/BMR-171086. | No African countries. |
| 21. | Kilic O, Maas M, Verhagen E, Zwerver J, Gouttebarge V. Incidence, aetiology and prevention of musculoskeletal injuries in volleyball: A systematic review of the literature. Eur J Sport Sci. 2017 Jul;17(6):765-793. doi: 10.1080/17461391.2017.1306114. | No meta-analysis. |
| 22. | Kilic Ö, Van Os V, Kemler E, Barendrecht M, Gouttebarge V. The 'Sequence of Prevention' for musculoskeletal injuries among recreational basketballers: a systematic review of the scientific literature. Phys Sportsmed. 2018 May;46(2):197-212. doi: 10.1080/00913847.2018.1424496. | No meta-analysis. |
| 23. | Kuo YL, Lee LL. Prevalence and risk factors associated with spinal pain in adolescent computer users: a systematic review. JBI Libr Syst Rev. 2012;10(45):2906-2943. doi: 10.11124/jbisrir-2012-26. | No meta-analysis. |
| 24. | Liao ZW, Le C, Kynes JM, Niconchuk JA, Pinto E, Laferriere HE, Walters CB. Paediatric chronic pain prevalence in low- and middle-income countries: A systematic review and meta-analysis. EClinicalMedicine. 2022 Feb 12;45:101296. doi: 10.1016/j.eclinm.2022.101296. | No specific meta-analyses for African countries.  African countries were jointly meta-analyzed with countries from other continents (e.g., Europe). |
| 25. | Louw QA, Morris LD, Grimmer-Somers K. The prevalence of low back pain in Africa: a systematic review. BMC Musculoskelet Disord. 2007 Nov 1;8:105. doi: 10.1186/1471-2474-8-105. | No meta-analysis. |
| 26. | Louw QA, Manilall J, Grimmer KA. Epidemiology of knee injuries among adolescents: a systematic review. Br J Sports Med. 2008 Jan;42(1):2-10. doi: 10.1136/bjsm.2007.035360. | No meta-analysis. |
| 27. | Migliorini F, Eschweiler J, Niewiera M, El Mansy Y, Tingart M, Rath B. Better outcomes with patellar resurfacing during primary total knee arthroplasty: a meta-analysis study. Arch Orthop Trauma Surg. 2019 Oct;139(10):1445-1454. doi: 10.1007/s00402-019-03246-z. | No specific meta-analyses for African countries. |
| 28. | Minervini G, Franco R, Marrapodi MM, Ronsivalle V, Shapira I, Cicciù M. Prevalence of temporomandibular disorders in subjects affected by Parkinson disease: A systematic review and metanalysis. J Oral Rehabil. 2023 Sep;50(9):877-885. doi: 10.1111/joor.13496. | No African countries. |
| 29. | Mnguni N, Olivier B, Mosselson J, Mudzi W. Prevalence of concurrent headache and temporomandibular disorders: a systematic review protocol. JBI Evid Synth. 2021 Jan;19(1):263-269. doi: 10.11124/JBISRIR-D-19-00255. | Protocol. |
| 30. | Mukhtar NB, Ibrahim AA, Mohammed J. Prevalence of neck pain and its associated factors in Africa: a systematic review and meta-analysis protocol. BMJ Open. 2023 Sep 18;13(9):e074219. doi: 10.1136/bmjopen-2023-074219. | Protocol. |
| 31. | Ndong A, Tendeng JN, Diallo AC, Diao ML, Sow O, Mawuli SD, Kalli M, Harissou A, Choua O, Doumga AD, Togo AP, Seck M, Ka I, Touré AO, Diop B, Ba PA, Diop PS, Cissé M, Sani R, Konaté I. Adult groin hernia surgery in sub-Saharan Africa: a 20-year systematic review and meta-analysis. Hernia. 2023 Feb;27(1):157-172. doi: 10.1007/s10029-022-02669-9. | No specific meta-analyses for musculoskeletal pain conditions. |
| 32. | Parker R, Stein DJ, Jelsma J. Pain in people living with HIV/AIDS: a systematic review. J Int AIDS Soc. 2014 Feb 18;17(1):18719. doi: 10.7448/IAS.17.1.18719. | No specific meta-analyses for African countries. |
| 33. | Pentapati KC, Yeturu SK, Siddiq H. Global and regional estimates of dental pain among children and adolescents-systematic review and meta-analysis. Eur Arch Paediatr Dent. 2021 Feb;22(1):1-12. doi: 10.1007/s40368-020-00545-7. | No specific meta-analyses for musculoskeletal pain conditions. |
| 34. | Rudan I, Sidhu S, Papana A, Meng SJ, Xin-Wei Y, Wang W, Campbell-Page RM, Demaio AR, Nair H, Sridhar D, Theodoratou E, Dowman B, Adeloye D, Majeed A, Car J, Campbell H, Wang W, Chan KY; Global Health Epidemiology Reference Group (GHERG). Prevalence of rheumatoid arthritis in low- and middle-income countries: A systematic review and analysis. J Glob Health. 2015 Jun;5(1):010409. doi: 10.7189/jogh.05.010409. | No specific meta-analyses for African countries.  Two studies were included but no formal meta-analysis was conducted for African countries. |
| 35. | Salari N, Sadeghi N, Hosseinian-Far A, Hasheminezhad R, Khazaie H, Shohaimi S, Mohammadi M. Prevalence of sleep disturbance in patients with ankylosing spondylitis: a systematic review and meta-analysis. Adv Rheumatol. 2023 Jul 19;63(1):33. doi: 10.1186/s42358-023-00315-1. | No objective of this study. |
| 36. | Stolwijk C, van Onna M, Boonen A, van Tubergen A. Global Prevalence of Spondyloarthritis: A Systematic Review and Meta-Regression Analysis. Arthritis Care Res (Hoboken). 2016 Sep;68(9):1320-31. doi: 10.1002/acr.22831. | No population of interest. |
| 37. | Stubbs B, Vancampfort D, Veronese N, Thompson T, Fornaro M, Schofield P, Solmi M, Mugisha J, Carvalho AF, Koyanagi A. Depression and pain: primary data and meta-analysis among 237 952 people across 47 low- and middle-income countries. Psychol Med. 2017 Dec;47(16):2906-2917. doi: 10.1017/S0033291717001477. | No specific meta-analyses for musculoskeletal pain conditions. |
| 38. | Stubbs B, Vancampfort D, Thompson T, Veronese N, Carvalho AF, Solmi M, Mugisha J, Schofield P, Matthew Prina A, Smith L, Koyanagi A. Pain and severe sleep disturbance in the general population: Primary data and meta-analysis from 240,820 people across 45 low- and middle-income countries. Gen Hosp Psychiatry. 2018 Jul-Aug;53:52-58. doi: 10.1016/j.genhosppsych.2018.05.006. | No specific meta-analyses for musculoskeletal pain conditions. |
| 39. | Usenbo A, Kramer V, Young T, Musekiwa A. Prevalence of Arthritis in Africa: A Systematic Review and Meta-Analysis. PLoS One. 2015 Aug 4;10(8):e0133858. doi: 10.1371/journal.pone.0133858. | No population of interest. |
| 40. | Wall J, Meehan WP 3rd, Trompeter K, Gissane C, Mockler D, van Dyk N, Wilson F. Incidence, prevalence and risk factors for low back pain in adolescent athletes: a systematic review and meta-analysis. Br J Sports Med. 2022 Nov;56(22):1299-1306. doi: 10.1136/bjsports-2021-104749. | No African countries. |
| 41. | Xu YW, Cheng AS, Li-Tsang CW. Prevalence and risk factors of work-related musculoskeletal disorders in the catering industry: a systematic review. Work. 2013;44(2):107-16. doi: 10.3233/WOR-2012-1375. | No African countries. |
| 42. | Yahaya I, Wright T, Babatunde OO, Corp N, Helliwell T, Dikomitis L, Mallen CD. Prevalence of osteoarthritis in lower middle- and low-income countries: a systematic review and meta-analysis. Rheumatol Int. 2021 Jul;41(7):1221-1231. doi: 10.1007/s00296-021-04838-y. | No population of interest. |
| 43. | Yusuf M, Gebrye T, Fatoye F. Burden of musculoskeletal-related disorders resulting from non-fatal road traffic collisions in Africa: a protocol of a systematic review. BMJ Open. 2019 Oct 28;9(10):e032687. doi: 10.1136/bmjopen-2019-032687. | Protocol. |
| 44. | Zhang L, Shen B, Liu S. Rheumatoid arthritis is associated with negatively variable impacts on domains of sleep disturbances: evidence from a systematic review and meta-analysis. Psychol Health Med. 2021 Mar;26(3):267-277. doi: 10.1080/13548506.2020.1764597. | No objective of this study. |

Manual searches:

Found in: Essouma M, Nkeck JR, Endomba FT, Bigna JJ, Ralandison S. Epidemiology of rheumatoid arthritis in sub-Saharan Africa: a systematic review and meta-analysis protocol. Syst Rev. 2020 Apr 17;9(1):81. doi: 10.1186/s13643-020-01342-5

- Essouma M, Noubiap JJ, Singwe-Ngandeu M, Hachulla E. Epidemiology of Idiopathic Inflammatory Myopathies in Africa: A Contemporary Systematic Review. J Clin Rheumatol. 2022 Mar 1;28(2):e552-e562. doi: 10.1097/RHU.0000000000001736. No meta-analysis.
- Essouma M, Noubiap JJ, Singwe-Ngandeu M, Hachulla E. Epidemiology of Sjögren Syndrome in Africa: A Scoping Review. J Clin Rheumatol. 2022 Jan 1;28(1):e240-e244. doi: 10.1097/RHU.0000000000001708. No meta-analysis.
- Essouma M, Nkeck JR, Endomba FT, Bigna JJ, Singwe-Ngandeu M, Hachulla E. Systemic lupus erythematosus in Native sub-Saharan Africans: A systematic review and meta-analysis. J Autoimmun. 2020 Jan;106:102348. doi: 10.1016/j.jaut.2019.102348. No objective of this study.
- Adelowo, O., Mody, G.M., Tikly, M. et al. Rheumatic diseases in Africa. Nat Rev Rheumatol 17, 363–374 (2021). <https://doi.org/10.1038/s41584-021-00603-4>. No meta-analysis.
