## Supplementary material for "THE PREVALENCE OF MUSCULOSKELETAL PAIN IN AFRICA: AN OVERVIEW OF SYSTEMATIC REVIEWS WITH META-ANALYSIS INCLUDING MORE THAN 100 DISTINCT PRIMARY STUDIES": Overlap LBP

**Supplementary file 3.** Matrix of evidence and the corrected covered area (CCA) calculation for low back pain.

| Number of studies without accounting for duplicates (r) | References of primary research | Systematic reviews where appear primary research including primary research duplicates (N) |
| --- | --- | --- |
| 1. | Abebaw T-A, Weldegebriel MK, Gebremichael B, Abaerei AA. Prevalence and associated factors of low back pain among teachers working at governmental primary schools in Addis Ababa, Ethiopia: a cross sectional study. Biomed J. 2018;1(6):3. | - Jegnie et al. 2021 - Mengistu et al. 2021 - Tesfaye et al. 2023 |
| 2. | Abebe AD, E. M. Gebrehiwot, S. Lema, and T. W. Abebe, “Prevalence of low back pain and associated risk factors among Adama Hospital Medical College Staff, Ethiopia,” European Journal of Preventive Medicine, vol. 3, no. 6, pp. 188–192, 2015. | - Jegnie et al. 2021 |
| 3. | Abiodun-Solanke I, Agbaje J, Ajayi D, Arotiba J. Prevalence of neck and back pain among dentists and dental auxillaries in south western Nigeria. Afr J Med Med Sci. 2010;39:137–42. | - Morris et al. 2018 |
| 4. | Adedoyin R, Idowu B, Adagunodo R, Ooyomi A, Idowu P. Musculoskeletal pain associated with the use of computer systems in Nigeria. Technol Health Care. 2005;13:125–30 | - Morris et al. 2018 |
| 5. | Adegoke B, Akodu A, Oyeyemi A. Work-related musculoskeletal disorders among Nigerian physiotherapists. BMC Musculoskel Disord. 2008;9:112. | - Morris et al. 2018 |
| 6. | Adegoke B, Odole A, Adeyinka A. Adolescent low back pain among secondary school students in Ibadan, Nigeria. Afr Health Sci. 2015;15:429–37 | - Morris et al. 2018 |
| 7. | Akinbo S, Odebiyi D, Osasan A. Characteristics of back pain among commercial drivers and motorcyclists in Lagos, Nigeria. West Afr J Med. 2008;27(2):85–9. | - Morris et al. 2018 |
| 8. | Akinpelu A, Oyewole O, Hammed G, Gbiri C. Prevalence of low back pain among adolescent students in a Nigerian Urban Community. AJPARS. 2013;5(1&2):29–34. | - Morris et al. 2018 |
| 9. | Akodu A, Taiwo A, Jimoh O. Prevalence of low back pain among traffic wardens in Lagos state. Nigeria AJPARS. 2014;6(1 & 2):37–41. | - Morris et al. 2018 |
| 10. | Akodu A, Okafor U, Adebayo A. Prevalence of low back pain among filling stations attendants in Lagos, Southwest Nigeria Afr J Biomed Res. 2016;19: 109–15. | - Morris et al. 2018 |
| 11. | Assefa H, Prevalence and Risk Factors of Low Back Pain in Nurses Working at Tikur Anbessa Specialized Hospital and Zewditu Memorial Hospital, Addis Ababa, Ethiopia, Addis Ababa University, Addis Ababa, Ethiopia, 2017 | - Jegnie et al. 2021 |
| 12. | Assefa T, Prevalnce of Work Related Lower Back Pain and Assocaiated Factors Among Welders in Selected Metal and Engineering Industries in Addis Ababa and Surrounding Towns, Addis Ababa University, Addis Ababa, Ethiopia, 2017 | - Jegnie et al. 2021 |
| 13. | Ayanniyi O, Sanya A, Ogunlade S, Oni-Orisan M. Prevalence and pattern of back pain among pregnant women attending ante-natal clinics in selected health care facilities. Afr J Biomed Res. 2006;9:149–56. | - Morris et al. 2018 |
| 14. | Ayanniyi O, Mbada C, Muolokwu C. Prevalence and profile of back pain in Nigerian adolescents. Med Princ Pract. 2011;20:368–73. | - Morris et al. 2018 |
| 15. | Bedru W, Self-Reported Work Related Musculoskeletal Disorders and Determinant Factors of Female Beauty Salon Hair Dressers, Addis Ababa University, Addis Ababa, Ethiopia, 2016. | - Jegnie et al. 2021 |
| 16. | Bejia I, Abid N, Salem K, Letaief M, Younes M, Touzi M, et al. Low back pain in a cohort of 622 Tunisian schoolchildren and adolescents: an epidemiological study. Eur Spine J. 2005;14:331–336 (a). | - Morris et al. 2018 |
| 17. | Bejia I, Younes M, Jamila H, Khalfallah T, Salem K, Touzi M, et al. Prevalence and factors associated with low back pain among hospital staff. Joint Bone Spine. 2005;72:254–259 (b). | - Morris et al. 2018 |
| 18. | Belay M, Worku A, Gebrie S, Wamisho B. Epidemiology of Low Back Pain among Nurses Working in Public Hospitals of Addis Ababa, Ethiopia East & Central African Journal of Surgery. 2016:21(1):113-31. | - Jegnie et al. 2021 - Kasa et al. 2020 - Morris et al. 2018 |
| 19. | Beyen TK, Mengestu MY, Zele YT. Low back pain and associated factors among teachers in Gondar Town, North Gondar, Amhara Region, Ethiopia. Occup Med Health Af. 2013;1(5):1–8. | - Jegnie et al. 2021 - Tesfaye et al. 2023 |
| 20. | Bezzaoucha A. Descriptive epidemiology of low-back pain in Algiers. Rev Rhum Mal Ostéoartic. 1992;59(2):121–4. | - Morris et al. 2018 |
| 21. | Bio F, Sadhra S, Jackson C, Burge P. Low back pain in underground gold miners in Ghana. Ghana Med J. 2007;41(1):21–5. | - Morris et al. 2018 |
| 22. | Birabi B, Dienye P, Ndukwu G. Prevalence of low back pain among peasant farmers in a rural community in south South Nigeria. Rural Remote Health. 2012;12:1920. | - Morris et al. 2018 |
| 23. | Booyens S, van Wyk P, Postma T. Musculoskeletal disorders amongst practising south African oral hygienists. SADJ. 2009;64(9):400–3. | - Morris et al. 2018 |
| 24. | Botha P, Chikte U, Barrie R, Esterhuizen T. Self-reported musculoskeletal pain among dentists in South Africa: a 12-month prevalence study. SADJ. 2014;69(5):208–13. | - Morris et al. 2018 |
| 25. | Boughattas W, Maalel O, El MM, Bougmiza I. Low Back pain among nurses: prevalence , and occupational risk factors. Occup Dis Environ Med. 2017;5: 26–37. https://doi.org/10.1016/j.annrmp.2007.05.009 | - Kasa et al. 2020 |
| 26. | Boukerma Z, Behlouli AL, Reggad M. Epidemiology of low back pain among nurses of the hospital of Sétif (Algeria). BMJ Occopational Environ Med. 2014;71(1):1 –6 A | - Kasa et al. 2020 |
| 27. | Chandeu M. Work-related low Back pain among clinical nurses in Tanzania: University of the Western Cape; 2008. | - Kasa et al. 2020 |
| 28. | Chiwaridzo M, Naidoo N. Prevalence and associated characteristics of recurrent non-specific low back pain in Zimbabwean adolescents: a cross-sectional study. BMC Musculoskelet Disord. 2014;15:381. | - Morris et al. 2018 |
| 29. | Chiwaridzo M, Makotore V, Dambi JM, Munambah N, Mhlanga M. Work - related musculoskeletal disorders among registered general nurses : a case of a large central hospital in Harare , Zimbabwe. BMC Res Notes. 2018; 11(315):1–7. https://doi.org/10.1186/s13104-018-3412-8. | - Kasa et al. 2020 |
| 30. | Dagne D, Abebe SM, Getachew A. Work-related musculoskeletal disorders and associated factors among bank workers in Addis Ababa, Ethiopia: a cross-sectional study. Environ Health Prev Med. 2020;25:33-38. | - Mengistu et al. 2021 |
| 31. | Desai F, Ellapen T, van Heerden H. The point prevalence of work-related musculoskeletal pain among general surgeons in KwaZulu-Natal, South Africa. Ergonomics SA. 2012;24(2):18–30. | - Morris et al. 2018 |
| 32. | El-Soud A, El-Najjar A, EL-Fattah N and Hassan A. Prevalence of low back pain in working nurses in Zagazig University hospitals: and epidemiological study. Egyptian Rheumatology and Rehabilitation. 2014;41:109–15 | - Morris et al. 2018 |
| 33. | Erick PN, Smith DR. Low back pain among school teachers in Botswana, prevalence and risk factors. BMC Musculoskelet Disord. 2014;15(1):1–13. | - Morris et al. 2018 - Tesfaye et al. 2023 |
| 34. | Etana G, Prevalence of Work Related Musculoskeletal Disorder and Associated Factors Among Bank Staff in Jimma City, Southwest Ethiopia, Jimma University, Jimma, Ethiopia, 2019. | - Jegnie et al. 2021 |
| 35. | Fabunmi A, ABa S, Odunaiya N. Prevalence of low back pain among peasant farmers in a rural community in south-West Nigeria. Afr J Med Med Sci. 2005;34(3):259–162. | - Morris et al. 2018 |
| 36. | Fahmy VF, Momen MAMT, Mostafa NS, Elawady MY. Prevalence, risk factors and quality of life impact of work-related musculoskeletal disorders among school teachers in Cairo, Egypt. BMC Public Health. 2022;22(1):1–17. | - Tesfaye et al. 2023 |
| 37. | Fanta M, Alagaw A, Kejela G, Tunje A. Low back pain and associated factors among civil service sectors office workers in southern Ethiopia. Int J Occup Saf Health. 2020;10:53-63 | - Mengistu et al. 2021 |
| 38. | Galukande M, Muwazi S, Mugisa D. Aetiology of low back pain in Mulago hospital, Uganda. Afr Health Sci. 2005;5:164–7. | - Morris et al. 2018 |
| 39. | Gebreyesus T, Weldemariam S, Fasika S, Abebe E, Kife M. Prevalence and associated factors of low back pain among school teachers in Mekelle City, Northern Ethiopia, 2016: a cross sectional study. World J Phys Med Rehab. 2019;1:1006. | - Tesfaye et al. 2023 |
| 40. | Girma Z, Assessing the Prevalence of Work Related Musculoskeletal Disorders and Associated Factors Among Workers in Selected Garments in Addis Ababa, Ethiopia, Addis Ababa University, Addis Ababa, Ethiopia, 2016. | - Jegnie et al. 2021 |
| 41. | Govender S. Low back pain in the nursing profession-a pilot study. SAOJ. 2004:7–13 | - Morris et al. 2018 |
| 42. | Hailu W, Getahun M, Mohammed A, et al. Assessment of back pain and disability status among automotive industry workers, in Ethiopia. Int J Sci Rep. 2020;6:301 | - Jegnie et al. 2021 - Mengistu et al. 2021 |
| 43. | Harris I. Prevalence of low back pain in cricketers-an undergraduate epidemiological study. Physiotherapy. 1993;49:65–6. | - Morris et al. 2018 |
| 44. | Henok A, Bekele T. Prevalence of musculoskeletal pain and factors associated with kyphosis among pedestrian back-loading women in selected towns of Bench Maji zone, Southwest Ethiopia. Ethiop J Health Dev. 2017;31:103-109. | - Mengistu et al. 2021 |
| 45. | Hill A, Darko R, Seffah J, Adanu R, Anarfi J, Duda R. Health of urban Ghanaian women as identified by the Women’s health study of Accra. Int J Gynaecol Obstet. 2007;99:150–6. | - Morris et al. 2018 |
| 46. | Himalowa S, Frantz J. The effect of occupationally-related low back pain on functional activities among male manual workers in a construction company in cape town, South Africa. Occup Healt SA. 2012;18(5):28–32. | - Morris et al. 2018 |
| 47. | Igumbor E, Useh U, Madzivire D. An epidemiological study of work-related low back pain among physiotherapists in Zimbabwe. SAJP. 2003;59:7–14. | - Morris et al. 2018 |
| 48. | Isa U, Saminu A, Rufai Y. Prevalence of low back pain complaints among commercial motorcyclists in Kano, Northwest Nigeria. Nigerian Medical Practitioner. 2009;56:19–23. | - Morris et al. 2018 |
| 49. | Jordaan R, Kruger M, Stewart A, Becker P. The association between low back pain, gender and age in adolescents. SAJP. 2005;61:15–20 | - Morris et al. 2018 |
| 50. | Kebede A, Abebe SM, Woldie H, Yenit MK: Low back pain and associated factors among primary school teachers in Mekele City, North Ethiopia: a cross-sectional study. Occupational therapy international 2019, 2019. | - Jegnie et al. 2021 - Mengistu et al. 2021 - Tesfaye et al. 2023 |
| 51. | Kibret AK, Fisseha Gebremeskel B, Embaye Gezae K, Solomon Tsegay G. Work-related musculoskeletal disorders and associated factors among bankers in Ethiopia, 2018. Pain Res Manag. 2020;2020:8735169. | - Mengistu et al. 2021 |
| 52. | Lela M. The relationship between physical activity and low Back pain among nurses in Kanombe military hospital: University of the Western Cape; 2010. | - Kasa et al. 2020 |
| 53. | Lette A, Hussen A, Kumbi M, Nuriye S, Lamore Y. Musculoskeletal pain and associated factors among building construction workers in southeastern Ethiopia. Ergo Int J. 2019;3:000214. | - Jegnie et al. 2021 - Mengistu et al. 2021 |
| 54. | Madiba S, Hoque M, Rakgase R. Musculoskeletal disorders among nurses in high acuity areas in a tertiary hospital in South Africa. Occup Healt SA. 2013;19(1):20–3. | - Morris et al. 2018 |
| 55. | Major-Helstoot M, Crous L, Grimmer-Somers K, Louw Q. Management of LBP at primary care level in South Africa: up to standards? Afr Health Sci. 2014;14:698–706. | - Morris et al. 2018 |
| 56. | Mbaye I, Fall M, Wone I, Dione P, Ouattara B, Sow M. Chronic low back pain in a Senegalese pubic transport’s company. Dakar Med. 2002;47(2):176–8. | - Morris et al. 2018 |
| 57. | Mekonnen TH. Work-related factors associated with low back pain among nurse professionals in east and west Wollega zones, western Ethiopia, 2017: a crosssectional study. Pain Ther. 2019;8:239-247. | - Jegnie et al. 2021 - Mengistu et al. 2021 |
| 58. | Mekonnen TH. The magnitude and factors associated with work-related back and lower extremity musculoskeletal disorders among barbers in Gondar town, northwest Ethiopia, 2017: a cross-sectional study. PLoS One. 2019;14:e0220035 | - Jegnie et al. 2021 - Mengistu et al. 2021 |
| 59. | Mekonnen TH, Kekeba GG, Azanaw J, Kabito GG. Prevalence and healthcare seeking practice of work-related musculoskeletal disorders among informal sectors of hairdressers in Ethiopia, 2019: findings from a cross-sectional study. BMC Public Health. 2020;20:718. | - Mengistu et al. 2021 |
| 60. | Melese H, Gebreyesus T, Alamer A, Berhe A. Prevalence and associated factors of musculoskeletal disorders among cleaners working at Mekelle University, Ethiopia. J Pain Res. 2020;13:2239-2246. | - Mengistu et al. 2021 |
| 61. | Mijena GF, B. Geda, M. Dheresa, and S. G. Fage, “Low back pain among nurses working at public hospitals in eastern Ethiopia,” Journal of Pain Research, vol. 13, pp. 1349–1357, 2020 | - Jegnie et al. 2021 |
| 62. | Mijiyawa M, Oniankitan O, Kolani B, Koriko T. Low back pain in hospital outpatients in Lome(Togo). Joint Bone Spine. 2000;67:533–8 | - Morris et al. 2018 |
| 63. | Mo E, Ebied E. Work-related musculoskeletal pain among primary school teachers: a recommended health promotion intervention for prevention and management. World J Nurs Sci. 2015;1(3):54–61. | - Tesfaye et al. 2023 |
| 64. | Muhammed AF, Awwal LM, Musa HA, Mustapha GA. Work-related risk factors for lower Back pain among nurses in Ahmadu Bello University teaching hospital ( ABUTH ), Zaria-Nigeria. IOSR J Nurs Heal Sci. 2015;4(3):20–5. | - Kasa et al. 2020 |
| 65. | Mulimba J. The problems of low back pain in Africa. East Afr Med J. 1990;67(4):250–3 | - Morris et al. 2018 |
| 66. | Munabi IG, Buwembo W, Kitara DL, Ochieng J, Mwaka ES. Musculoskeletal disorder risk factors among nursing professionals in low resource settings : a cross-sectional study in Uganda. BMC Nurs. 2014;13(7):1 –8. | - Kasa et al. 2020 |
| 67. | Mwaka E, Munabi I, Buwembo W, Kukkiriza J, Ochieng J. Musculoskeletal pain and school bag use: a cross-sectional study among Ugandan pupils. BMC Research Notes. 2014;7:222. | - Morris et al. 2018 |
| 68. | Mwangi A, Downing R, Elias HE. Low back pain among primary school teachers in Rural Kenya: Prevalence and contributing factors. African J Primary Health Care Family Med. 2019;11(1):1–7. | - Tesfaye et al. 2023 |
| 69. | Naidoo R, Coopoo Y. The health and fitness profiles of nurses in KwaZulu-Natal. Curationis. 2007;30(2):66–73 | - Morris et al. 2018 |
| 70. | Ndonye NA, Matara NJ, Muriithi IA: Predictors of work-related musculoskeletal disorders among primary school teachers in Machakos County, Kenya. Int J Ind Ergon 2019. | - Tesfaye et al. 2023 |
| 71. | Nilahi CD: Work-related lower back pain among primary school teachers in Dar es Salaam, Tanzania. 2014. | - Tesfaye et al. 2023 |
| 72. | Noorbhai M, Essack F, Thwala S, Ellapen T, van Heerden J. Prevalence of cricket-related musculoskeletal pain among adolescent cricketers in KwaZulu-Natal. SAJSM. 2012;24(1):3–9. | - Morris et al. 2018 |
| 73. | Odebiyi D, Ogwezi D, Adegoke B. The prevalence of low back pain in commercial motor drivers and private automobile drivers. Nig J Med Rehabil. 2007;21(1 & 2):21-4 | - Morris et al. 2018 |
| 74. | Odebiyi D, Akanle O, Akinbo S, Balogun S. Prevalence and impact of workrelated musculoskeletal disorders on job performance of call center operators in Nigeria. Int J Occup Environ Med. 2016;7:98–106. | - Morris et al. 2018 |
| 75. | Ogunbode A, Adebusoye L, Alonge T, Ogunbode A. Prevalence of low back pain and associated risk factors amongst adult patients presenting to a Nigerian family practice clinic, a hospital-based study. Afr J Prim Health Care. 2012;5(1):1–8. | - Morris et al. 2018 |
| 76. | Olana AT. Occupational risk factors of low back pain among ammunition engineering industry in West Shoa Zone, Ethiopia, 2017. J Med Physiol Biophys. 2018;45:31-36 | - Mengistu et al. 2021 |
| 77. | Omokhodion F, Umar U, Ogunnowo B. Prevalence of low back pain among staff in a rural hospital in Nigeria. Occup Med. 2000;50:107–10. | - Kasa et al. 2020 - Morris et al. 2018 |
| 78. | Omokodion F. Low back pain in a rural community in south West Nigeria. West Afr J Med. 2002;2:87–90. | - Morris et al. 2018 |
| 79. | Omokodion F, Sanya A. Risk factors for low back pain among office workers in Ibadan, Southwest Nigeria Short report. Occ Med. 2003;53:287–9. | - Morris et al. 2018 |
| 80. | Omokodion F. Low back pain in an urban population in Southwest Nigeria. Trop Dr. 2004;34:17–20. | - Morris et al. 2018 |
| 81. | Ouédraogo D, Ouédraogo V, Ouedraogo L, Kinda M, Tiéno H, Zoungrana E, et al. Prevalence and risk factors associated with low back pain among hospital staff in Ouagadougou (Burkina Faso). Med Trop. 2010;70:277–80. | - Morris et al. 2018 |
| 82. | Prista A, Balague F, Nordin M, Skovron M. Low back pain in Mozambican adolescents. Eur Spine J. 2004;13:341–5. | - Morris et al. 2018 |
| 83. | Regassa TM, Lema TB, Garmomsa GN. Work related musculoskeletal disorders and associated factors among nurses working in Jimma Zone public hospitals, South West Ethiopia. Occup Med Health Aff. 2018;6:2. | - Jegnie et al. 2021 - Mengistu et al. 2021 |
| 84. | Rufa’i A, Sa’idu I, Ahmad R, Elmi O, Aliyu S, Jarere A, et al. Prevalence and risk factors for low back pain among professional drivers in Kano. Nigeria: Archives of Environmental and Occupational Health; 2013. [Epub ahead of print]. | - Morris et al. 2018 |
| 85. | Sa’idu I, Utti V, Jaiyesimi A, Rufa’i A, Maduagwu S, Onuwe H, et al. Prevalence of musculoskeletal injuries among factory workers in Kano metropolis, Nigeria. Int J Occup Saf Ergon. 2011;17:99–102. | - Morris et al. 2018 |
| 86. | Sanya A, Ogwumike O. Low back pain prevalence amongst industrial workers in the private sector in Oyo state, Nigeria. Afr J Med Med Sci. 2005;34:245–9. | - Morris et al. 2018 |
| 87. | Schierhout G, Myers J, Bridger R. Musculoskeletal pain and workplace ergonomic stressors in manufacturing industry in South Africa. Int J Ind Ergon. 1993;12:3–11. | - Morris et al. 2018 |
| 88. | Sikiru L, Shmaila H. Prevalence and risk factors of low back pain among nurses in Africa: Nigerian and Ethiopian specialized hospitals survey study. East Afr J Pub Health. 2009;6(1):22–6. | - Jegnie et al. 2021 - Kasa et al. 2020 - Morris et al. 2018 |
| 89. | Sikiru L, Hanifa S. Prevalence and risk factors of low back pain among nurses in a typical Nigerian hospital. Afr Health Sci. 2010;10(1):26–30. | - Kasa et al. 2020 - Morris et al. 2018 |
| 90. | Sumaila FG, Mayana KI, Bello B, Sharif AM. Prevalence of low back pain and back ergonomics awareness among teachers of selected secondary schools in Kano Metropolis. Int J Physiother. 2015;2(6):999–1005. | - Tesfaye et al. 2023 |
| 91. | Tafese A, “Occupational risk factors of low back pain among ammunition engineering industry in West Shoa Zone, Ethiopia, 2017,” Journal of Medicine, Physiology and Biophysics, vol. 45, 2018. | - Jegnie et al. 2021 |
| 92. | Tafese A, Kebede G, Shibru A, Benti T. Work-Related low back pain among sewing machine operators of garment industry: Galan City Oromia region, Ethiopia. Int J Occup Hyg. 2018;10:1-6. | - Jegnie et al. 2021 - Mengistu et al. 2021 |
| 93. | Tamene A, Mulugeta H, Ashenafi T, Thygerson SM. Musculoskeletal disorders and associated factors among vehicle repair workers in Hawassa city, southern Ethiopia. J Environ Public Health. 2020;2020:9472357. | - Mengistu et al. 2021 |
| 94. | Teklu S, Assessment of Prevalence and Associated Factors of Work Related Musculoskeletal Disorders Among Cobble Stone Workers in Addis Ababa, Ethiopia, Addis Ababa University, Addis Ababa, Ethiopia, 2017. | - Jegnie et al. 2021 |
| 95. | Tella B, Akinbo S, Asafa S, Gbiri C. Prevalence and impacts of low back pain among peasant farmers in south-West Nigeria. Int J Occup Med Environ Health. 2013;26:621–7. | - Morris et al. 2018 |
| 96. | Thembelihle. Prevalence of Low Back Pain Amongst Nurses at Edendale Hospital A dissertation submitted to the Department of Public Health Medicine Nelson R . Mandela School of Medicine Durban , South Africa In partial fulfilment of the requirements for the Master in Pu. University of KwaZulu-Natal; 2010. | - Kasa et al. 2020 |
| 97. | Thembelihle D, AV& SK. Prevalence and factors associated with low back pain among nurses at a regional hospital in KwaZulu-Natal, South Africa. Heal SA Gesondheid.. 2014;1–6. Available from: http://www.hsag.org.za. Accessed 25 Sept 2018. | - Kasa et al. 2020 |
| 98. | Tinubu B, Mbada C, Oyeyemi A, Fabunmi A. Work-related musculoskeletal disorders among nurses in Ibadan, south-West Nigeria: a cross-sectional survey. BMC Musculoskel Disord. 2010;11:12. | - Kasa et al. 2020 - Morris et al. 2018 |
| 99. | Tolera ST, Kabeto SK. Occupational-Related musculoskeletal disorders and associated factors among beauty salon workers, Adama Town, south-eastern Ethiopia, 2018. J Ergo. 2020;9:257. | - Mengistu et al. 2021 |
| 100. | Triki M, Koubaa A, Masmoudi L, Fellman N, Tabka Z. Prevalence and risk factors of low back pain among undergraduate students of a sports and physical education institute in Tunisia. Libyan J Med. 2015;10:26802. | - Morris et al. 2018 |
| 101. | Van Vuuren B, Becker P, Van Heerden H, Zinzen E, Meeusen R. Lower back problems and occupational risk factors in a south African steel industry. Am J Ind Med. 2005;47:45–457 | - Morris et al. 2018 |
| 102. | Van Vuuren B, Zinzen E, Van Heerden H, Becker P, Meeusen R. Psychosocial factors related to lower back problems in a south African manganese industry. J Occup Rehabil. 2005;15:215–25. | - Morris et al. 2018 |
| 103. | Vincent-Onabajo G, Nweze E, Kachalla G, Ali M, Usman A, Alhaji M, Umeonwuka C. Prevalence of Low Back Pain among Undergraduate Physiotherapy Students in Nigeria. Pain Res Treat. 2016;123038:4 | - Morris et al. 2018 |
| 104. | Wami SD, G. Abere, A. Dessie, and D. Getachew, “Workrelated risk factors and the prevalence of low back pain among low wage workers: results from a cross-sectional study,” BMC Public Health, vol. 19, no. 1, p. 1072, 2019. | - Jegnie et al. 2021 |
| 105. | Wanamo ME, Abaya SW, Aschalew AB. Prevalence and risk factors for low back pain (LBP) among taxi drivers in Addis Ababa, Ethiopia: a community based cross-sectional study. Ethiop J Health Dev. 2017;31:244-250. | - Jegnie et al. 2021 - Mengistu et al. 2021 |
| 106. | Worku Z. Prevalence of low back pain in Lesotho mothers. JMPT. 2000;23:147–54 | - Morris et al. 2018 |
| 107. | Yehualaw W, Assessment of Self-Reported Work Related Low Back Pain and Associated Factors Among Nurses Working in Intensive Care Unit (ICU) at Public and Private Hospitals, Addis Ababa, Ethiopia, Addis Ababa University, Addis Ababa, Ethiopia, 2017 | - Jegnie et al. 2021 |
| 108. | Yitayeh A, S. Mekonnen, S. Fasika, and M. Gizachew, “Annual prevalence of self-reported work related musculoskeletal disorders and associated factors among nurses working at Gondar Town Governmental Health Institutions, Northwest Ethiopia,” Emergency Medicine, vol. 5, no. 227, p. 2, 2015. | - Jegnie et al. 2021 |
| 109. | Yosef T, Belachew A, Tefera Y. Magnitude and contributing factors of low back pain among long distance truck drivers at Modjo dry port, Ethiopia: a cross-sectional study. J Environ Public Health. Published online September 22, 2019. doi:10.1155/2019/6793090 | - Mengistu et al. 2021 |

| CCA = | N-r | = | 129-109 | 20 | = 0.046 = 5% |
| --- | --- | --- | --- | --- | --- |
|  | rc-r |  | 545-109 | 436 |  |

Note: N is the total number of included publications (including duplicates) in the meta-analyses (the sum of all checked boxes in the citation matrix, as shown in Supplementary File 3). Furthermore, r is the number of studies without accounting for duplicates. Finally, c is the number of systematic reviews included in the evidence matrix (k = 5). CCA = corrected covered area.

Note: Kasa et al. 2020 did not report adequately the list of references. Therefore, we only could extract for the overlap calculation 14 out of 18 studies (Lamina et al. appeared twice. For that reason, we did not count 19 studies in this section, here). The following studies were not found: Asmare et al. 2015; Mengestie et al. 2016; Betty 2015; Amany et al. 2014.
