## Supplementary material for "THE PREVALENCE OF MUSCULOSKELETAL PAIN IN AFRICA: AN OVERVIEW OF SYSTEMATIC REVIEWS WITH META-ANALYSIS INCLUDING MORE THAN 100 DISTINCT PRIMARY STUDIES": PRIOR checklist

(Gates M, Gates A, Pieper D, et al. Reporting guideline for overviews of reviews of healthcare interventions: development of the PRIOR statement. *BMJ* 2022;378:e070849. doi:10.1136/bmj-2022-070849.)

| **Section**  Topic | **#** | **Item** | **Location reported** |
| --- | --- | --- | --- |
| **TITLE** | | |  |
| Title | 1 | Identify the report as an overview of reviews. | 1 |
| **ABSTRACT** | | |  |
| Abstract | 2 | Provide a comprehensive and accurate summary of the purpose, methods, and results of the overview of reviews. | 3 |
| **INTRODUCTION** | | |  |
| Rationale | 3 | Describe the rationale for conducting the overview of reviews in the context of existing knowledge. | 5 |
| Objectives | 4 | Provide an explicit statement of the objective(s) or question(s) addressed by the overview of reviews. | 6 |
| **METHODS** | | |  |
| Eligibility criteria | 5a | Specify the inclusion and exclusion criteria for the overview of reviews. If supplemental primary studies were included, this should be stated, with a rationale. | 7-8 |
|  | 5b | Specify the definition of ‘systematic review’ as used in the inclusion criteria for the overview of reviews. | 8 |
| Information sources | 6 | Specify all databases, registers, websites, organizations, reference lists, and other sources searched or consulted to identify systematic reviews and supplemental primary studies (if included).  Specify the date when each source was last searched or consulted. | 7 |
| Search strategy | 7 | Present the full search strategies for all databases, registers and websites, such that they could be reproduced. Describe any search filters and limits applied. | 7 and Suppl File 1 |
| Selection process | 8a | Describe the methods used to decide whether a systematic review or supplemental primary study (if included) met the inclusion criteria of the overview of reviews. | 8 |
|  | 8b | Describe how overlap in the populations, interventions, comparators, and/or outcomes of systematic reviews was identified and managed during study selection. | 10 |
| Data collection process | 9a | Describe the methods used to collect data from reports. | 9 |
|  | 9b | If applicable, describe the methods used to identify and manage primary study overlap at the level  of the comparison and outcome during data collection. For each outcome, specify the method used to illustrate and/or quantify the degree of primary study overlap across systematic reviews. | 10 |
|  | 9c | If applicable, specify the methods used to manage discrepant data across systematic reviews during data collection. | NO |
| Data items | 10 | List and define all variables and outcomes for which data were sought. Describe any assumptions made and/or measures taken to identify and clarify missing or unclear information. | 8 |
| Risk of bias assessment | 11a | Describe the methods used to *assess* risk of bias or methodological quality of the included systematic reviews. | 9 |
|  | 11b | Describe the methods used to *collect* data on (from the systematic reviews) and/or *assess* the risk of bias of the primary studies included in the systematic reviews. Provide a justification for instances where flawed, incomplete, or missing assessments are identified but not re-assessed. | NO |
|  | 11c | Describe the methods used to *assess* the risk of bias of supplemental primary studies (if included). | Not applicable |
| Synthesis methods | 12a | Describe the methods used to summarize or synthesize results and provide a rationale for the choice(s). | 10 No rationale but justification was provided |
|  | 12b | Describe any methods used to explore possible causes of heterogeneity among results. | Not applicable |
|  | 12c | Describe any sensitivity analyses conducted to assess the robustness of the synthesized results. | Not applicable |
| Reporting bias assessment | 13 | Describe the methods used to *collect* data on (from the systematic reviews) and/or *assess* the risk of bias due to missing results in a summary or synthesis (arising from reporting biases at the levels of the systematic reviews, primary studies, and supplemental primary studies, if included). | NO |
| Certainty assessment | 14 | Describe the methods used to *collect* data on (from the systematic reviews) and/or *assess* certainty (or confidence) in the body of evidence for an outcome. | Not applicable |
| **RESULTS** | | |  |
| Systematic review and supplemental primary study selection | 15a | Describe the results of the search and selection process, including the number of records screened, assessed for eligibility, and included in the overview of reviews, ideally with a flow diagram. | 11 and Figure 1 |
|  | 15b | Provide a list of studies that might appear to meet the inclusion criteria, but were excluded, with the main reason for exclusion. | Suppl File 2 |

| **Section**  Topic | **#** | **Item** | **Location**  **reported** |
| --- | --- | --- | --- |
| Characteristics of systematic reviews and  supplemental primary studies | 16 | Cite each included systematic review and supplemental primary study (if included) and present its characteristics. | Table 1 |
| Primary study overlap | 17 | Describe the extent of primary study overlap across the included systematic reviews. | 12 and Suppl Files 3-5 |
| Risk of bias in systematic reviews, primary studies, and  supplemental primary studies | 18a | Present assessments of risk of bias or methodological quality for each included systematic review. | 12 and  Table 2 |
|  | 18b | Present assessments (*collected* from systematic reviews or *assessed* anew) of the risk of bias of  the primary studies included in the systematic reviews. | NO |
|  | 18c | Present assessments of the risk of bias of supplemental primary studies (if included). | Not applicable |
| Summary or synthesis of results | 19a | For all outcomes, summarize the evidence from the systematic reviews and supplemental primary studies (if included). If meta-analyses were done, present for each the summary estimate and its  precision and measures of statistical heterogeneity. If comparing groups, describe the direction of the effect. | 12-14 and Table 1 |
|  | 19b | If meta-analyses were done, present results of all investigations of possible causes of  heterogeneity. | Not applicable |
|  | 19c | If meta-analyses were done, present results of all sensitivity analyses conducted to assess the  robustness of synthesized results. | Not applicable |
| Reporting biases | 20 | Present assessments (*collected* from systematic reviews and/or *assessed* anew) of the risk of bias due to missing primary studies, analyses, or results in a summary or synthesis (arising from reporting biases at the levels of the systematic reviews, primary studies, and supplemental primary  studies, if included) for each summary or synthesis assessed. | NO |
| Certainty of  evidence | 21 | Present assessments (*collected* or *assessed* anew) of certainty (or confidence) in the body of  evidence for each outcome. | Table 1 (GRADE was not used for any review) |
| **DISCUSSION** | | |  |
| Discussion | 22a | Summarize the main findings, including any discrepancies in findings across the included systematic reviews and supplemental primary studies (if included). | 14-15 |
|  | 22b | Provide a general interpretation of the results in the context of other evidence. | NO |
|  | 22c | Discuss any limitations of the evidence from systematic reviews, their primary studies, and supplemental primary studies (if included) included in the overview of reviews. Discuss any  limitations of the overview of reviews methods used. | 18 |
|  | 22d | Discuss implications for practice, policy, and future research (both systematic reviews and  primary research). Consider the relevance of the findings to the end users of the overview of reviews, e.g., healthcare providers, policymakers, patients, among others. | 15-17 |
| **OTHER INFORMATION** | | |  |
| Registration and protocol | 23a | Provide registration information for the overview of reviews, including register name and registration number, or state that the overview of reviews was not registered. | 7 and abstract |
|  | 23b | Indicate where the overview of reviews protocol can be accessed, or state that a protocol was not  prepared. | 7 and abstract |
|  | 23c | Describe and explain any amendments to information provided at registration or in the protocol.  Indicate the stage of the overview of reviews at which amendments were made. | 7 |
| Support | 24 | Describe sources of financial or non-financial support for the overview of reviews, and the role of  the funders or sponsors in the overview of reviews. | Title page and abstract |
| Competing  interests | 25 | Declare any competing interests of the overview of reviews' authors. | Title page |
| Author information | 26a | Provide contact information for the corresponding author. | Title page |
|  | 26b | Describe the contributions of individual authors and identify the guarantor of the overview of  reviews. | Title page: partially |
| Availability of data and other materials | 27 | Report which of the following are available, where they can be found, and under which conditions they may be accessed: template data collection forms; data collected from included systematic  reviews and supplemental primary studies; analytic code; any other materials used in the overview of reviews. | Not applicable |
